# Decision support for preventing elective surgery cancellations: cost-sensitive risk ranking with cross-site validation in the NHS

**DOI:** 10.64898/2026.07.03.26357241

**Authors:** Hassan Chizari, Noel Peters, Benjamin Lin, Fahimeh Malekinezhad, Mark Pietroni

**Affiliations:** University of Gloucestershire; Gloucestershire Hospitals NHS Foundation Trust

**Keywords:** elective surgery, no-show, cost-sensitive learning, domain shift, XGBoost, NHS, decision support

## Abstract

Elective surgery late cancellations and “did not attend” (LCDNA) events waste theatre capacity, lengthen waiting lists, and impose avoidable costs on NHS Trusts. We present a decision-support approach that ranks upcoming elective procedures by expected cancellation cost and supports capacity-constrained outreach by selecting the highest-risk Top-*K* cases for intervention. Using cost-sensitive learning and a clinically grounded cost model, the policy reduces expected cost from approximately £103 per case under business-as-usual to £77.08 per case in a hospital-holdout (cross-site) evaluation designed to mimic deployment to a new hospital. In a complementary time-forward evaluation, representing prospective use within the same service environment, expected cost falls further to £70.97 per case. The £6.11 per-case difference between the two regimes highlights the added uncertainty introduced by cross-site operational shift and supports a conservative roll-out with local calibration and monitoring. Explainability analyses suggest that booking-to-procedure lead time, specialty or service line, calendar effects, and prior cancellation history are the strongest drivers of prediction, helping to inform tiered intervention workflows that prioritise near-term bookings and use model–pathway mismatches as an audit signal. Overall, the framework turns predictive performance into practical, capacity-aware policy guidance for reducing avoidable cancellations while supporting safe and equitable implementation.

## 1. Introduction

The National Health Service (NHS) in the United Kingdom is facing persistent pressure in elective surgical care, with long waiting lists and substantial operational inefficiencies Svenson (2023). Late Cancellations and Did Not Attend (LCDNA) cases are a particularly costly part of this problem because they occur too close to the scheduled procedure to allow effective list reorganisation, wasting theatre capacity and staff time while delaying care for other patients Central London Patient Safety Research Collaboration (2024); Getting It Right First Time (GIRFT) (2024). At Gloucestershire Hospitals NHS Foundation Trust, the setting for this study, annual losses linked to cancelled lists and early finishes are substantial, with 3,369 on-the-day cancellations recorded in FY24/25 alone.

Traditional responses to theatre disruption are often local, manual, and reactive. National guidance increasingly emphasises digital visibility, systematic monitoring, and learning from trends rather than isolated cancellation events Scottish Health Technologies Group (SHTG) (2023); NHS England (2025). Machine learning is attractive in this setting because it can combine patient, scheduling, and operational factors to support earlier risk identification. However, most prior cancellation-prediction work has focused on outpatient or primary care settings, with limited attention to NHS elective surgery LCDNA and limited emphasis on deployment-relevant cost-sensitive decision making Tuan et al. (2025).

Our aim is not simply to identify cases at higher risk of LCDNA, but to support a practical policy for allocating limited pre-operative contact capacity. Here, “intervention” refers to a high-touch pre-operative support bundle (typically led by nurses or administrators), not an automated SMS reminder; it may include telephone confirmation, barrier resolution, checking completion of pre-assessment, escalation to pre-assessment, and proactive rescheduling to protect theatre capacity.

**Contributions of this paper are:** (i) a cost-sensitive formulation of LCDNA risk ranking that links predictions to an explicit expected-cost objective and demonstrates substantial savings versus business-as-usual; (ii) a practical, capacity-aware top-*K* intervention policy that converts model scores into a workload-compatible contact/support list; (iii) a dual-regime validation design that separately stress-tests temporal drift (time-forward) and cross-site operational shift (hospital-holdout), translating methodological rigour into deployment-relevant savings guarantees; (iv) interpretable analyses that translate lead-time and service-line effects into prescriptive tiered workflows and pathway-audit prompts; and (v) a deployment governance roadmap covering monitoring, local calibration, and fairness checks across demographic subgroups.

Prior work on missed appointments and elective-care access suggests that attendance and cancellation risk may vary with age, sex, deprivation, transport barriers, digital access, and caring or work constraints; for that reason, fairness monitoring is treated here as a deployment requirement rather than an optional extra.

The paper proceeds as follows. Section 2 sets out the background and rationale, including the NHS waiting-list context, theatre utilisation challenges, and related work on cancellation prediction. Section 3 describes the methodology, including the dataset, feature engineering, model development, and evaluation strategy. Section 4 reports the results, including model performance, cost comparisons with baseline strategies, and feature-importance analyses. Section 5 discusses the findings, operational implications, limitations, and future work. Section 6 concludes with recommendations for deployment. Section 7 summarises the main points for policy makers.

## 2. Background and Rationale

### 2.1. The NHS Waiting List crisis

NHS elective care remains under severe pressure, with 6.2 million patients waiting in England alone (7.4 million across the UK), 3.2 million waiting more than 18 weeks, and approximately 194,000 waiting more than a year Svenson (2023); Salisbury et al. (2023); Mahase (2024). This backlog, intensified by the COVID-19 pandemic, has increased pressure on hospitals to improve operating-theatre utilisation, reduce avoidable disruption, and manage waiting lists more effectively Darzi (2024); The King’s Fund (2024).

### 2.2. Theatre Utilisation and Cancellations

Operating theatres are among the NHS’s most valuable and costly assets, so improving their utilisation is central to elective recovery. National policy responses such as surgical hubs aim to increase throughput through ring-fenced elective pathways and ambitious theatre utilisation targets Royal College of Surgeons of England (2024); Poole et al. (2025). At the same time, the GIRFT PACE audit reported a 9.9% cancellation rate for elective procedures within 24 hours of surgery across participating NHS Trusts, with booking and scheduling issues, acute medical problems, and patient non-attendance among the main contributors Central London Patient Safety Research Collaboration (2024); Getting It Right First Time (GIRFT) (2024). Short-notice patient-initiated cancellation is therefore an operationally important target for intervention.

### 2.3. Late Cancellation Cost

Late cancellation or non-attendance of elective surgery before or on the day of scheduled date results in substantial waste of operating theatre capacity. UK system-level estimates suggest that around 135,000 operations are cancelled on the day of surgery each year Centre for Perioperative Care (2024), at a cost of approximately £400 million in lost operating theatre time to the NHS Gillies et al. (2018), implying an average direct cost of nearly £3,000 per cancelled case Royal College of Anaesthetists (2024). Micro-costing studies from individual hospitals are consistent with this order of magnitude. Briggs (2019) report a typical running cost of about £1,200 per hour for an NHS operating theatre, while Baker et al. (2025) estimate that 7,405 minutes of theatre time lost to 94 same-day cancellations in a UK teaching hospital corresponded to £96,265 of wasted capacity (valuing theatre time at £13 per minute), i.e. roughly £1,000 per cancelled case. International data are similar: a Finnish university hospital reported a mean financial loss of Euro 2,459.91 for each elective day-of-surgery cancellation Turunen et al. (2018). NHS and European literature estimates for cancelled day-case or elective procedures typically range from £1,110 to £1,220 per cancellation. Taken together, these figures suggest that a per-cancellation cost in the range of £1,000–£3,000 is plausible for elective procedures that cannot be backfilled, so modelling a conservative cost of £1,000 per LCDNA event in our setting is likely to underestimate, rather than overstate, the true economic impact.

By contrast, the marginal cost of simple pre-emptive interventions to reduce LCDNA events is comparatively small. NHS England estimates that a missed general practice appointment costs around £30 NHS England (2019), while a local evaluation of hospital outpatient activity quotes average losses of £108–£160 per missed appointment, reflecting the full cost of staff time and overheads for a standard visit Shropshire Council (2016). Behavioural interventions that alter the content of SMS reminders can reduce non-attendance "at no additional cost" beyond an existing messaging infrastructure Hallsworth et al. (2015), and digital pre-operative triage pathways such as PRO-MAPP have been shown to be cost-saving overall, reducing preoperative costs by about £769 per hip or knee surgery patient compared with standard care Gleave (2023).

We set the intervention cost to *i* = *£*100 per contacted case to represent a high-touch, nurse- or administrator-led support bundle requiring staff time and overhead. This composite assumption combines: (1) SMS/phone reminder costs (consistent with prior evaluations Hallsworth et al. (2015) and Shropshire Council (2016)), (2) nurse review or virtual pre-operative clinic time (approximately 10–20 minutes at typical NHS banding rates) plus documentation and escalation where required, and (3) administrative overhead. The assumption is deliberately conservative. £100 is higher than the implied marginal cost of SMS-only strategies (which correspond to much smaller values and can be analysed directly in our framework in sensitivity analyses), yet remains an order of magnitude lower than the expected loss from an unfilled operating-theatre slot following LCDNA.

The resulting cost ratio *m/i* = 10 (where *m* = *£*1,000 and *i* = *£*100) is intended as a conservative estimate of the relative economic impact. It remains reasonable under plausible variation: even if the true missed-cancellation cost were as low as £750 (yielding *m/i* = 7.5) or as high as £1,250 (yielding *m/i* = 12.5), the qualitative conclusion that targeted intervention is cheaper than doing nothing still holds (see Section 4.3.2 for detailed sensitivity analysis). The £100 intervention cost is also conservative, since many reminder-based interventions have lower marginal costs, while the bundle assumed here (telephone/virtual contact plus reminders) is relatively resource-intensive and therefore acts as an upper-bound estimate.

### 2.4. Machine Learning (ML) in healthcare operations

Machine Learning (ML) automation in healthcare operations involves using technology to streamline and optimise processes, reducing the need for manual intervention. The benefits of using ML in healthcare environment include improved data compliance, reduced backlogs, and significant time savings. For example, automating the management of "Did Not Attend" (DNA) appointments helps trusts quickly identify and follow up with patients Dawoodbhoy et al. (2021); Wall (2024).

Primary care settings have been a common focus for cancellation prediction research. For instance, a large-scale study analysing over one million appointments across 15 family medicine clinics developed a personalised machine learning approach that integrated clinical, geosocial, and environmental data to predict both no-shows and late cancellations Tuan et al. (2025). In another study, Liu et al. (2019) developed a machine learning approach to predict cancellation of children’s surgery by mining patient-specific and contextual data, demonstrating the value of comprehensive feature engineering for surgical cancellation prediction. Luo et al. (2020) focused on identifying surgeries with high risk of cancellation, specifically addressing peri-operative cancellations and highlighting the importance of pre-operative risk assessment. Zhang et al. (2021) conducted a systematic exploration of key experimental factors in ML-based surgical cancellation prediction, examining how different modelling choices affect performance. Li et al. (2024) developed an ML-based approach to predict last-minute cancellation of paediatric day surgeries, achieving strong performance. Liu et al. (2021) explored paediatric surgery cancellation through geospatial analysis and machine learning of short-notice cancellations, demonstrating the value of incorporating geographic factors. Sardesai et al. (2024) conducted a systematic review of day-of-surgery cancellation prediction, providing a comprehensive overview of the field. These studies’ success in different settings provides a foundation for understanding how similar approaches might be applied to surgical contexts, though the different operational characteristics and patient populations suggest that specialised models may be necessary.

Across the surgical and appointment cancellation literature, evaluation is still dominated by internal random splits or cross-validation within a single setting, leaving an important gap around deployment generalisation. Prior studies often train and test on randomly sampled records from one organisation or time-mixed pool (for example, a 70/30 random split across two campuses within a single children’s hospital Liu et al. (2019), single-site 80/20 splits for institutional cancellations Luo et al. (2020); Zhang et al. (2021), and *k*-fold procedures where folds are drawn from the same overall distribution Liu et al. (2021); Tuan et al. (2025)), with performance reported mainly via AUC (or RMSE) rather than deployment-style testing. This leaves two related questions unresolved: *(i) time-forward validity*, because random splitting mixes past and future pathways and can mask temporal drift in booking practices, waiting-list pressures, and perioperative processes. Steyerberg and Harrell Jr (2015) argue strongly against random splits in medical prediction settings for this reason. *(ii) domain-shift validity*, because models trained and tested within one site (or closely related campuses) may not transfer well to new hospitals with different case-mix, specialty composition, and operational policies. Random splits can also increase the risk of information leakage, for example through repeated patients, near-duplicate episodes, or process-related variables appearing in both training and test sets, which can inflate apparent performance and overstate projected savings. In short, strong internal metrics in prior work do not fully answer the practical deployment question: whether the model and its implied intervention policy will remain reliable on *future theatre lists* and/or in a *different hospital environment*. Against that background, Gloucestershire Hospitals NHS Foundation Trust (GHNFT), working with the University of Gloucestershire (UOG), developed the machine learning approach examined in this paper.

## 3. Methodology

This section describes the methodological approach used in the study. It begins with the dataset and its main characteristics, then outlines model development, and finally explains model selection and evaluation.

### 3.1. Dataset

The dataset is an extract of elective surgical theatre scheduling records from Gloucestershire Hospitals NHS Foundation Trust, spanning five theatre sites: Cheltenham General Hospital (CGH), Gloucestershire Royal Hospital (GRH), the Trust’s community hospital sites (TWC and CIR), and Stroud General Hospital (STG). The extract covers procedures scheduled between 2024–04–09 and 2025–04–29 (procedure dates) with booking dates from 2024–04–02 to 2025–04–01.

After applying the inclusion and filtering rules described below, the analytical dataset contains *N* = 20,282 scheduled procedures from 17,763 unique patients. The binary target variable indicates whether the case resulted in a LCDNA or proceeding to operation. Overall LCDNA rate is 11.1% (2,244 LCDNA out of 20,282).

#### Potential leakage and exclusion principles

The upstream operational dataset contains fields that may only be known after the outcome occurs (e.g., actual theatre times, performed procedure fields, cancellation dates/codes). To prevent label leakage, we impose a *safe feature gate* in preprocessing. This removes 54 potential leakage columns in our implementation.

#### Patient-level identifier

This column is used only to enforce patient exclusivity across splits and folds; it is never used as a predictive feature.

#### Other cancellations

The database contains explicit columns for cancellation reason categories, which classify cancellations into four main types: Clinical Cancellation, List Reorganise, Non-Clinical Cancellation, and Patient Cancellation. LCDNA cases were defined as records where the cancellation reason code indicated Patient Cancellation (patient-initiated cancellations or no-shows). The database also contains explicit columns for cancellation timing, including an on-the-day cancellation indicator and the "within three days" cancellations. LCDNA cases were restricted to those where the cancellation occurred within three days of the scheduled procedure date (including on-the-day cancellations). Clinical Cancellations, List Reorganise cases, and Non-Clinical Cancellations were excluded from the LCDNA definition by filtering records based on the cancellation reason code, ensuring that only patient-initiated cancellations were included in this work.

### 3.2. Data Coding and Feature Engineering

Features are grouped into: (i) Patient attributes (e.g., age at admission, sex, encoded comorbidity indicators such as asthma/COPD/cancer flags including a IS Sum which has the total number of comorobities for a patient); (ii) Procedure and scheduling context (e.g., specialty, day-of-week indicators, public holiday proximity, booking-to-procedure lead-time and bucketed lead-time); and (iii) Patient history aggregates computed from prior records within the dataset (e.g., prior appearances, prior cancellations, and a smoothed cancellation rate). (iv) Geographic features which are based on postcode. Postcode is available only at outward-code granularity in this extract (e.g., first 3 characters). We therefore do not link to deprivation indices such as IMD and treat fine-grained geographic proxies (e.g., full postcode patterns) as out-of-scope for this study. Where present, distance-to-hospital fields were ablated and found not to improve cost consistently under domain shift. Figure 1 shows the percentage of LCDNA versus operations on some of the variables used in this research.

**Figure 1:**
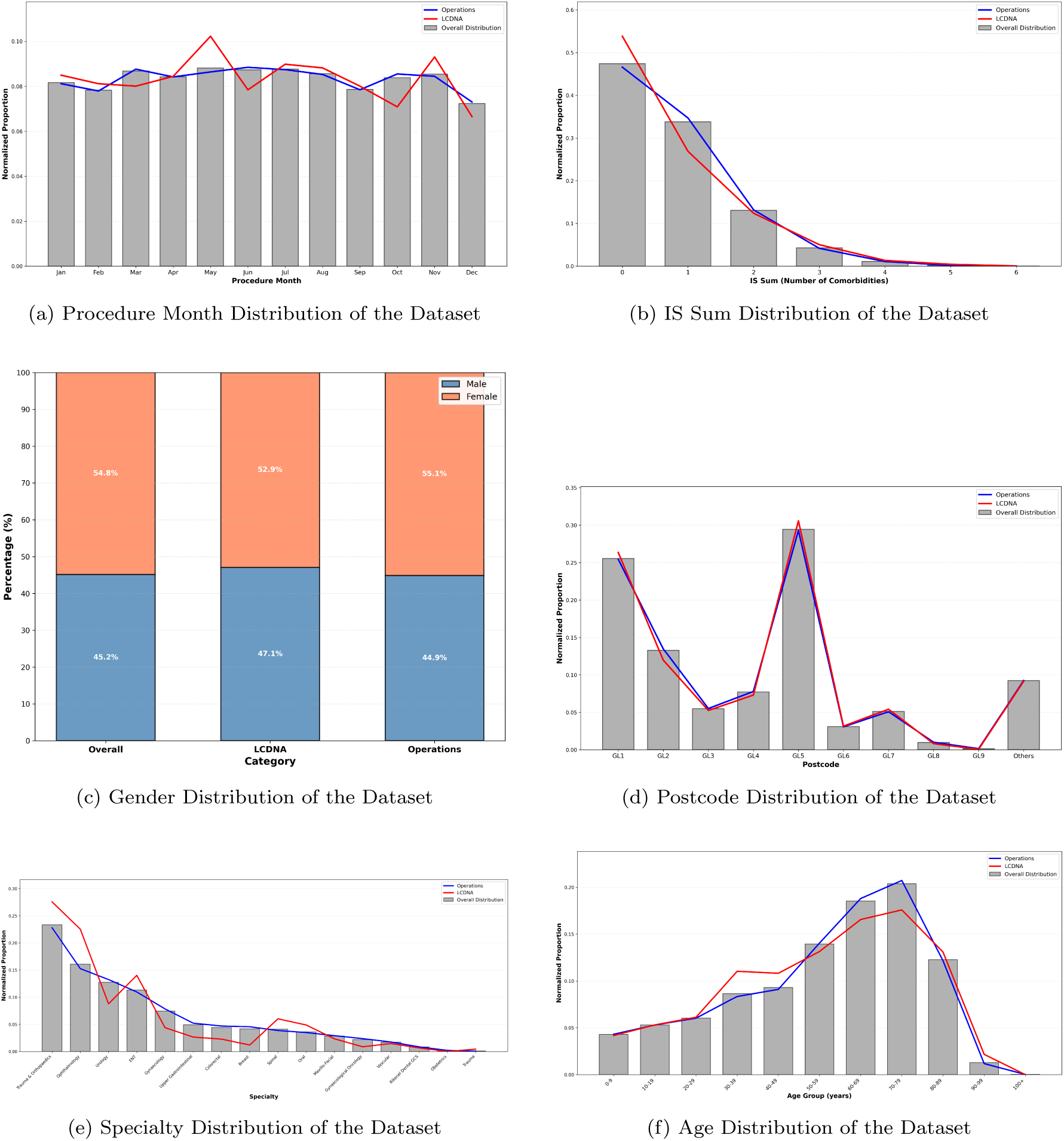
Distribution Analysis of Dataset Features

**Figure 2:**
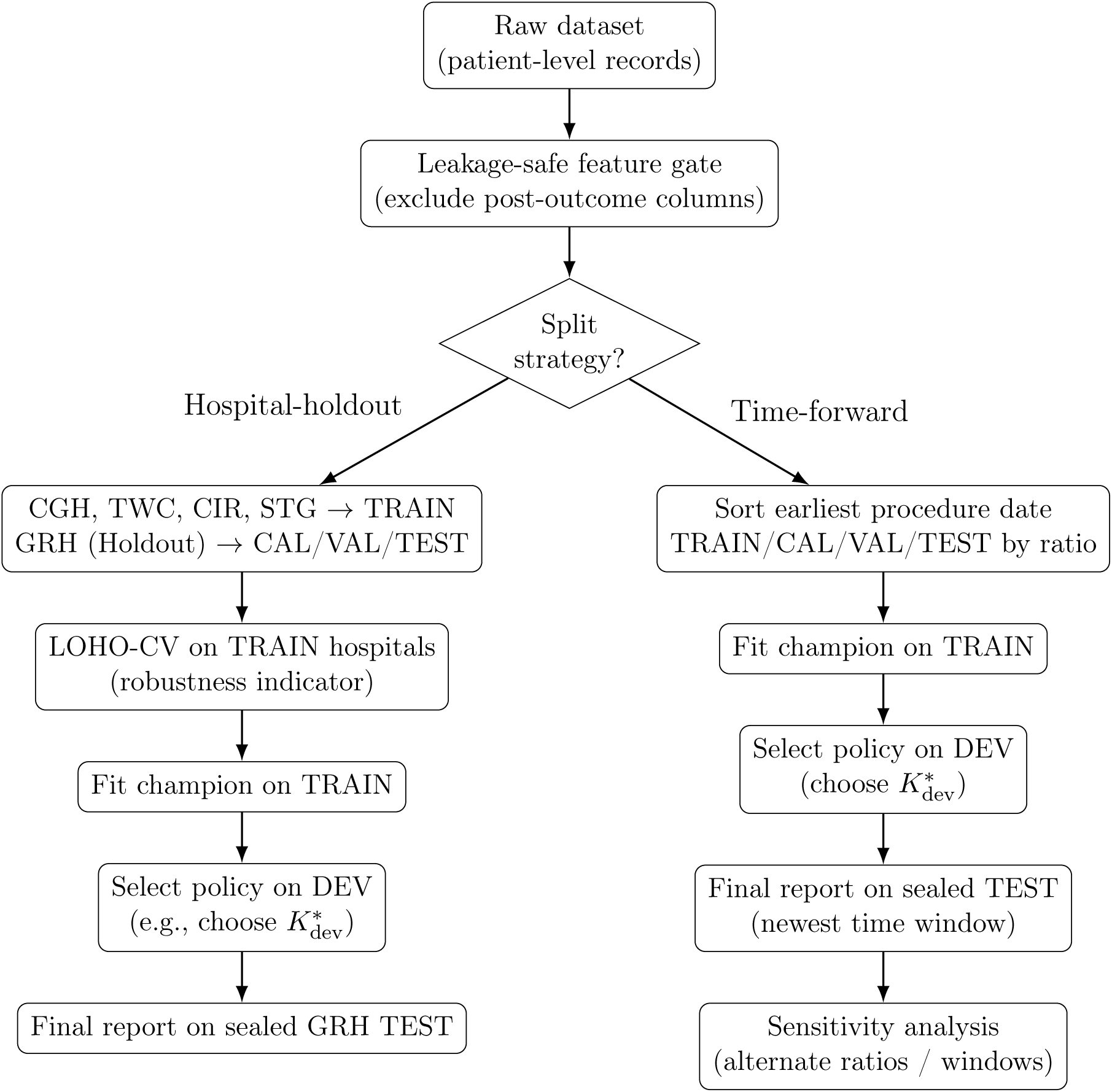
Evaluation workflow used in this study

**Figure 3:**
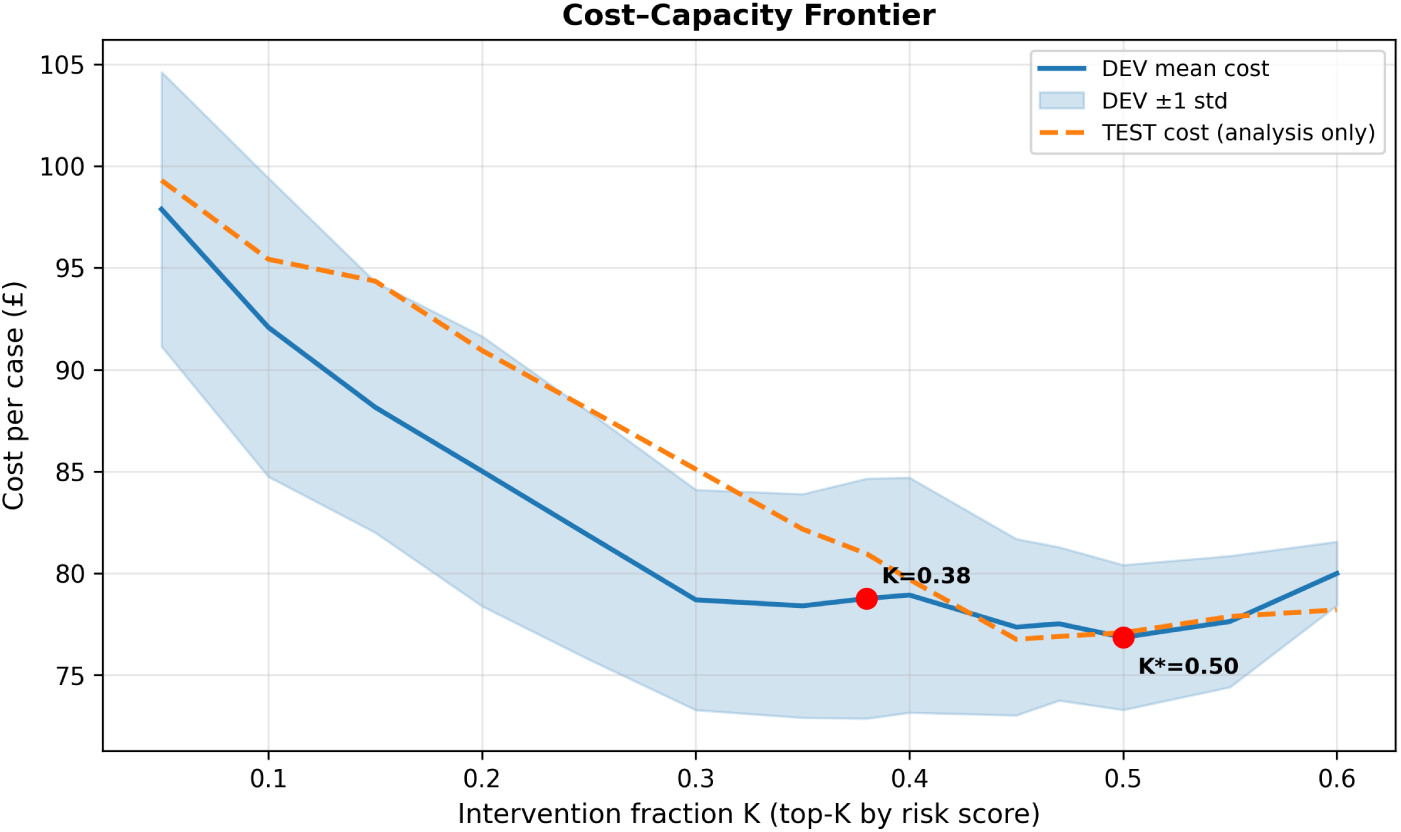
Cost–capacity frontier under a global top-*K* intervention policy. The TEST curve is shown for analysis only and was not used for model or policy selection.

**Figure 4:**
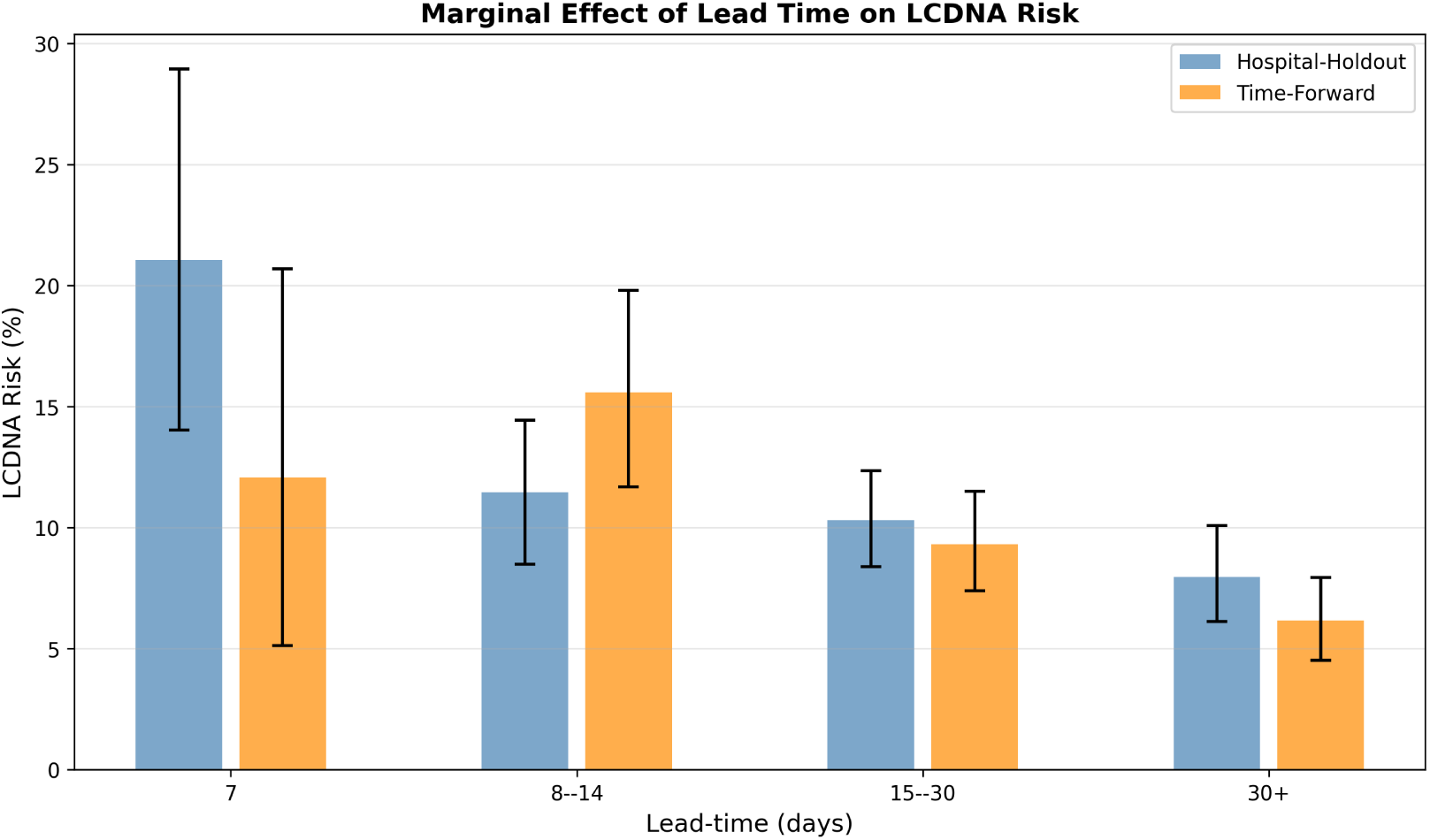
Marginal effect of lead time on LCDNA risk with bootstrap uncertainty: Hospital-Holdout vs Time-Forward models.

**Figure 5:**
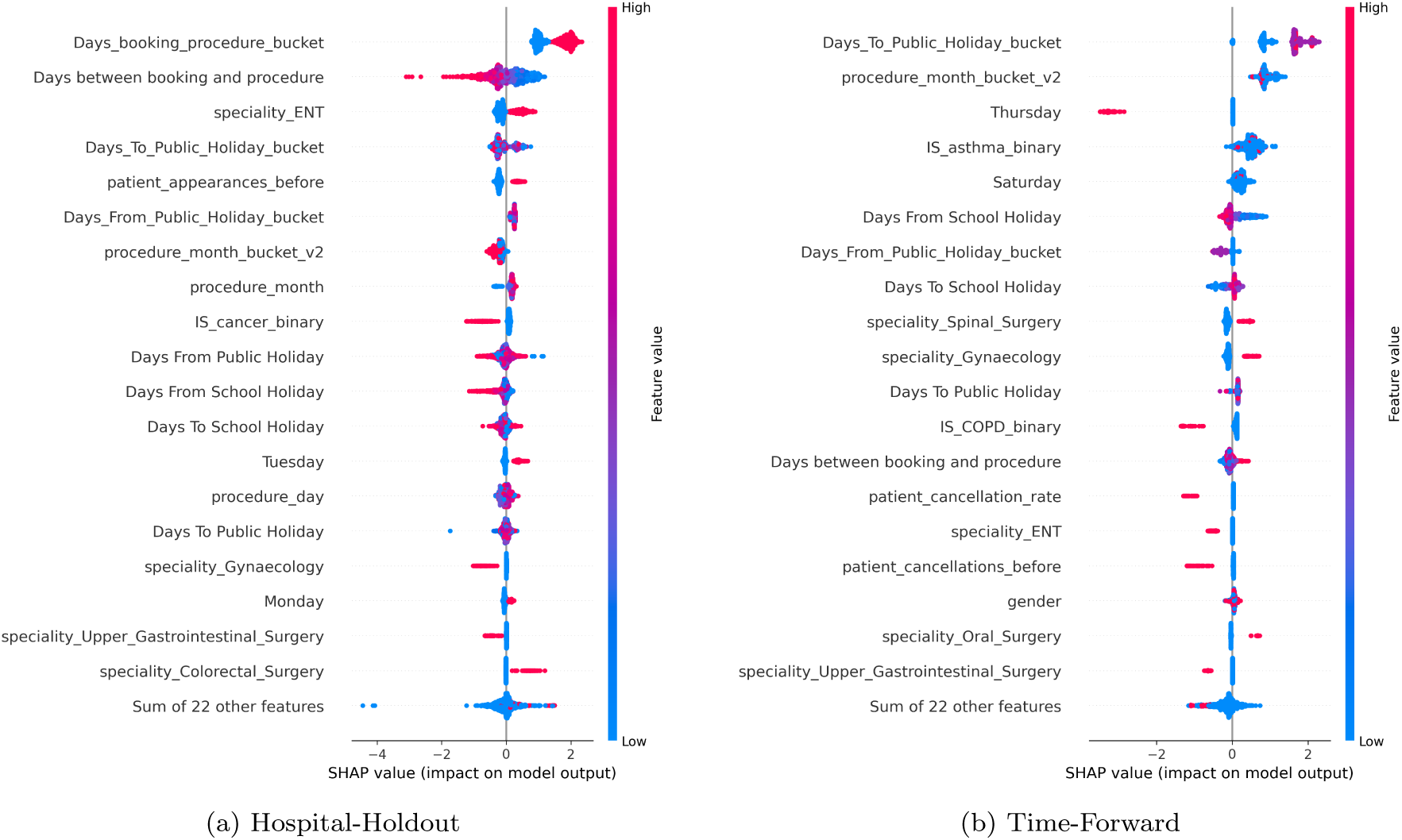
SHAP beeswarm plots showing feature impact distribution

### 3.3. Feature Engineering and Time Series Learning Methodology

Predicting LCDNA cases requires attention to both feature engineering and temporal structure, because outcomes depend on a mix of relatively stable patient characteristics and time-varying operational patterns.

Comprehensive feature engineering has been shown to improve healthcare LCDNA prediction, particularly when models combine heterogeneous clinical, geosocial, environmental, and temporal data into structured predictors Tuan et al. (2025). In line with prior evidence that schedule lead time is one of the strongest drivers of missed appointments and late cancellation risk Chiam et al. (2021); McMullen and Netland (2015); Mohammadi et al. (2018), we derived temporal, clinical, patient-history, and postcode-based geographic features from information available before the actionable window. Temporal validity was preserved by using only time-aware splitting and chronological validation, which provides more realistic deployment estimates than random record-level splitting in healthcare prediction settings Morid et al. (2023); Zhao et al. (2017); Steyerberg and Harrell Jr (2015).

Feature selection was iterative and decision-focused: we began with the full eligible feature set and removed features that did not contribute to operational performance, with the objective of retaining a compact set that reduced expected cost while remaining interpretable.

### 3.4. Model Development Strategy

#### 3.4.1. Time-Aware Data Splitting

To better reflect deployment conditions, we evaluate the approach under two complementary non-random regimes Steyerberg and Harrell Jr (2015):

##### Hospital-holdout (domain shift) evaluation (primary)

We designate one hospital site as a deployment target (GRH) and treat it as a holdout domain. Patients who have any procedures at non-holdout hospitals are assigned to Train set, ensuring that the holdout contains patients whose observed care pathway is exclusively within the target hospital. Holdout patients are then split chronologically by procedure date (by each patient’s earliest procedure date at the holdout hospital) into three disjoint subsets: CAL (policy calibration), VAL (model/policy selection robustness checks) and TEST (sealed final report). Patient-level exclusivity is enforced across all splits, with all procedures for each patient assigned to the split determined by their earliest procedure date.

To ensure strict domain separation and address potential concerns about cross-site patient contamination, we excluded all patients who had procedures at both GRH and train hospitals from the analysis. This exclusion removed 231 unique cross-site patients who collectively had 512 total procedures (247 at GRH and 265 at train hospitals). After this exclusion, using GRH as the deployment target, the training set contains *n* = 13, 040 procedures from 11, 338 unique patients, primarily at CGH (11, 571), TWC (881), CIR (388) and STG (200). The holdout hospital GRH is partitioned chronologically by procedure date into three roughly equal subsets: CAL (*n* = 2, 348; 2, 064 patients), VAL (*n* = 2, 239; 2, 064 patients) and TEST (*n* = 2, 142; 2, 066 patients), with LCDNA rates of 11.54%, 10.99% and 10.32% respectively. Patients are assigned to splits based on their earliest procedure date, with procedure date ranges (earliest per patient): CAL (2024-04-10 to 2024-08-27), VAL (2024-08-27 to 2024-12-23) and TEST (2024-12-23 to 2025-04-29). All procedure date ranges for GRH subsets are: CAL (2024-04-10 to 2025-04-29), VAL (2024-08-27 to 2025-04-28) and TEST (2024-12-23 to 2025-04-29).

##### Time-forward (deployment-like) evaluation (secondary)

We additionally perform a patient-level, chronological split based on each patient’s earliest procedure date, where older procedures form TRAIN and the most recent patients form TEST. This mirrors a “train on history, deploy on the future” workflow. It also provides a complementary view of temporal generalisation when the deployment domain is unchanged.

Using split ratios of 60/15/15/10, the resulting partitions contained: TRAIN 12,717 rows from 10,657 patients (ratio 0.1168; procedure dates 2024-04-09..2024-11-22), CAL 2,959 rows from 2,664 patients (ratio 0.1081; procedure dates 2024-11-22..2025-01-23), VAL 2,814 rows from 2,664 patients (ratio 0.0970; procedure dates 2025-01-23..2025-03-18), and TEST 1,791 rows from 1,778 patients (ratio 0.0927; procedure dates 2025-03-18..2025-04-29). All five hospitals were represented in each split; for example, TEST comprised CGH (1,036), GRH (638), TWC (56), CIR (49), and STG (12) cases.

##### Sealed test principle

In all experiments, TEST is used *once* for a final report. Any tuning of model hyperparameters or policy parameters (e.g., intervention budget) is performed using TRAIN and development data (CAL/VAL or DEV folds) only.

#### 3.4.2. Preprocessing Pipeline

A comprehensive preprocessing pipeline was applied to prepare the data for model training. Missing values were handled using median imputation for all numeric features, implemented through the *SimpleImputer* class from scikit-learn with the *strategy=’median’* parameter. Median imputation was chosen over mean imputation because it is more robust to outliers and preserves the distributional characteristics of the data. The imputation was fitted only on the training data, and the fitted *imputer* was then applied to the validation and test sets, ensuring that no information from future data leaked into the training process.

Feature scaling was performed using the *RobustScaler*, which standardises features by removing the median and scaling according to the interquartile range (IQR). RobustScaler was chosen over StandardScaler because it is less sensitive to outliers, which are common in healthcare data. The scaler was fitted only on the training data, and the fitted scaler was then applied to the validation and test sets, ensuring that no information from future data leaked into the training process.

#### 3.4.3. Models

The final modelling pipeline uses *gradient-boosted decision trees* as the primary learner, implemented via *XGBClassifier* Chen and Guestrin (2016). XGBoost performs well on heterogeneous tabular data, handles missingness robustly, supports monotonic regularisation, and produces risk rankings that suit a capacity-aware top-*K* intervention policy.

To test sensitivity to imbalance-handling choices, we also implemented the following alternative strategies for comparison (all trained on TRAIN only):

- **Scale-pos-weight only:** XGBoost with class weighting based on TRAIN prevalence.
- **No imbalance weighting:** XGBoost with *scale_pos_weight* =1.
- **ADASYN oversampling He et al. (2008):** XGBoost trained on ADASYN-resampled TRAIN at multiple target ratios.
- **Balanced bagging:** BalancedBaggingClassifier with an XGBoost base estimator (from the imbalanced-learn ecosystem) Lemaître et al. (2017).

Among these alternatives, the tuned XGBoost “champion” configuration (Section 4.1) was selected because it delivered the most robust cost performance under cross-hospital evaluation and remained stable across repeated random seeds.

#### 3.4.4. Class Imbalance Handling

LCDNA events are a minority class (overall prevalence ≈ 11%), which motivates explicit imbalance handling. We explored both algorithmic class weighting (`XGBoost scale_pos_weight`) and data-level resampling (ADASYN). However, our results show that heavy resampling does not consistently reduce expected operational cost under domain shift; in several configurations it increased false positives without commensurate reduction in false negatives. The final champion therefore uses class weighting and regularisation, and relies on a capacity-aware intervention policy (top-*K*) to balance recall against operational burden.

#### 3.4.5. Model Architecture and Training

All models are trained on TRAIN dataset only. Hyperparameters for the champion XGBoost model were tuned on development folds to minimise mean cost under a fixed top-*K* policy (Section 4.1.1). The best-performing hyperparameters were:

We further evaluated stability by repeating training under multiple random seeds (*n* = 100 seeds) and found low variance in both DEV and TEST cost: coefficient of variation (CV) was 1.52% for DEV and 1.86% for TEST, supporting reproducibility of the champion configuration.

#### 3.4.6. Cost-Based Threshold optimisation

Instead of optimising a classification threshold in isolation, we treat decision-making as a cost-sensitive intervention policy. Each case receives a predicted probability *p̂* of LCDNA, and the operational question is whether to *intervene* (e.g., reminder, confirmation, rescheduling, or clinical review) before surgery. We model two unit costs: (i) a missed LCDNA cost *m* = *£*1,000 (false negative), representing avoidable theatre under-utilisation and downstream disruption; and (ii) an intervention cost *i* = *£*100 applied whenever an intervention is triggered (true positive or false positive), representing staff time and communication overhead.

Given a confusion matrix (*TP, FP, FN, TN*) on a dataset of size *N*, the total and per-case costs are:

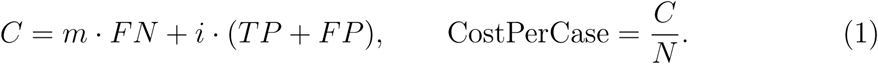

This formulation assumes *perfect intervention effectiveness*: true positives (TP) avoid the full avoidable cost *m* with no residual cost. This represents an upper-bound assumption on intervention success; in practice, interventions may have partial effectiveness (see Section 4.3.2 for comprehensive sensitivity analysis).

##### Capacity-aware top-*K* policy (primary)

Because hospitals have finite outreach capacity, the primary policy intervenes on the *K* · *N* cases with the highest predicted risk *p̂* for a chosen budget *K* ∈ (0, 1). This aligns model output directly with operational workload.

##### Selecting *K* on development data

We select *K* by minimising mean Cost-PerCase on rolling development folds (DEV) constructed from CAL+VAL, and we report a full cost–capacity frontier to support decision-making under different operational budgets. TEST remains sealed and is used only for the final report at the development-selected *K*.

##### Threshold-based policy (secondary)

For completeness, we also implemented cost-based threshold selection on CAL (predict LCDNA if *p̂* ≥ *t*) and evaluated it on VAL/TEST. However, the final pipeline and results reported in this paper use the capacity-aware top-*K* policy because it better matches real intervention constraints and yielded more stable performance under domain shift.

#### 3.4.7. Probability Calibration and Threshold Exploration

Because capacity-aware top-*K* policies depend on the *ranking* of predicted risk rather than absolute probability values, the final pipeline primarily relies on raw model scores. Nevertheless, we evaluated probability calibration methods (none vs Platt sigmoid Platt (1999) vs isotonic calibration Zadrozny and Elkan (2002)) on development folds. Calibration did not yield consistent improvements in expected cost and, in some settings, increased variance. Therefore, the chosen configuration uses uncalibrated XGBoost scores with a top-*K* intervention policy.

#### 3.4.8. Model Selection and Evaluation

##### Common protocol (applies to all split strategies)

Across all experiments, we (i) apply a leakage-safe feature gate (excluding columns that are only populated post-outcome), (ii) train candidate models using TRAIN only, (iii) select model/policy settings using development data only (CAL/VAL and/or rolling DEV folds), and (iv) report final performance on the sealed TEST split. Performance is measured using the cost-sensitive top-*K* intervention policy, where intervening on a case incurs a fixed intervention cost and missed cancellations incur a higher false-negative cost.

#### Path A. Hospital-holdout evaluation (cross-hospital generalisation)

To assess generalisation to a new site, we hold out an entire hospital (GRH) and train on the remaining hospitals. Patients with procedures at training hospitals are assigned to TRAIN; patients exclusively associated with GRH are assigned to the holdout pool and then split chronologically into CAL/VAL/TEST using each patient’s earliest procedure date, with strict patient-level exclusivity across all splits. Model selection is performed on development data only. In addition, we assess cross-hospital robustness within the training hospitals using leave-one-hospital-out cross-validation (LOHO-CV): for each training hospital *h*, a model is trained on TRAIN\*h* and evaluated on *h*; LOHO-CV mean cost is used as an auxiliary robustness indicator. The final champion is then refit on all TRAIN hospitals and evaluated once on the sealed GRH TEST split. Policy setting for capacity is performed using development data (CAL/VAL and rolling folds), yielding a recommended operating point 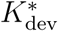 (selected without reference to TEST). No direct clinical intervention was delivered; the top-*K* policy was evaluated retrospectively.

##### B. Time-forward evaluation (deployment-like temporal generalisation)

To approximate deployment in a stable hospital system over time, we use a patient-level chronological split based on each patient’s earliest `operation_date when missing`). Patients are sorted chronologically and assigned to TRAIN / CAL / VAL / TEST according to a fixed ratio (default 60 / 15 / 15 / 10), enforcing strict patient-level exclusivity across splits. The model is trained on the historical TRAIN segment, policy parameters (including the recommended intervention fraction 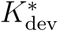) are selected using development segments (CAL/VAL and/or rolling DEV folds), and final performance is reported once on the sealed TEST window (the newest time period). We further perform sensitivity analysis by varying the time-forward split ratios and test-window length, while preserving the sealed-test protocol.

## 4. Results

### 4.1. Primary Evaluation: Hospital-Holdout (GRH) with Capacity-Aware Intervention

We report results for the hospital-holdout setting where GRH is treated as the deployment target. The model is trained on TRAIN (multi-site) and the intervention policy is selected on development data (CAL/VAL; DEV folds). Table 2 summarises the headline performance across four scenarios: *K*=0 represents the current “do nothing” baseline where no interventions are made; *K*=1 represents intervening on all cases; the historical baseline policy budget *K*=0.38; and the development-selected recommendation *K*^⋆^=0.50. For the sealed TEST set, the corresponding cost estimates are £103.17 [91.00, 116.77] for *K*=0, £80.95 [73.15, 90.39] for *K*=0.38, £77.08 [71.00, 84.84] for *K*^⋆^=0.50, and £100.00 [100.00, 100.00] for *K*=1, using 95% Wilson intervals on the observed TEST counts. Compared to the do-nothing baseline (*K*=0), the optimal policy (*K*^⋆^=0.50) achieves a cost reduction of £26.10 per case (25.3% savings), while the historical baseline (*K*=0.38) achieves £22.22 per case (21.5% savings). Compared to the intervene-all strategy (*K*=1), both *K*=0.38 and *K*^⋆^=0.50 provide savings of £19.05 (19.0%) and £22.92 (22.9%) per case respectively, demonstrating that targeted interventions based on predicted risk are more cost-effective than universal intervention.

**Table 1:**
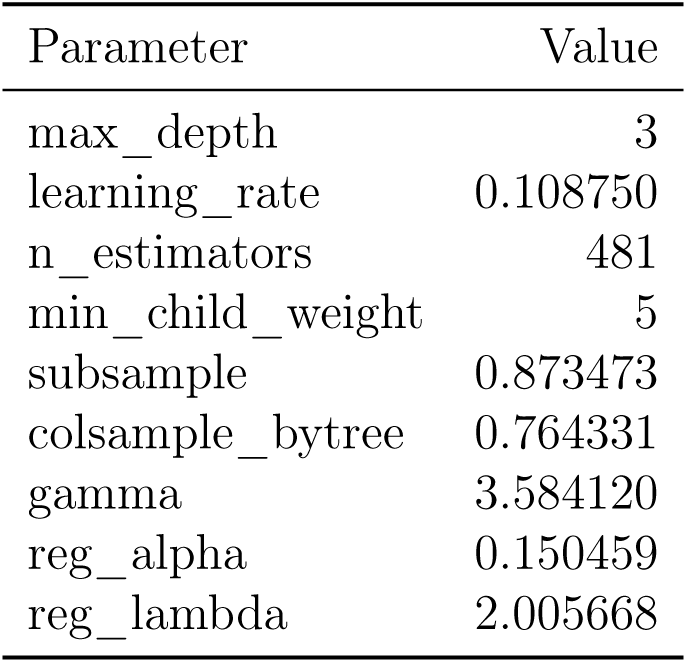
Champion XGBoost hyperparameters (mean-tuned configuration)

**Table 2:** Headline hospital-holdout results (GRH)

| $K$ | DEV<br>mean±std | TEST<br>cost [95% CI] | TP/FP/FN/TN | %int | int<br>cost | missed<br>cost |
| --- | --- | --- | --- | --- | --- | --- |
| 0.00 | 108.97±7.90 | 103.17 [91.00, 116.77] | 0/0/221/1921 | 0.00 | 0.00 | 103.17 |
| 0.38 | 78.76±5.90 | 80.95 [73.15, 90.39] | 129/685/92/1236 | 38.00 | 38.00 | 42.95 |
| 0.50 ($K^*$) | 76.85±3.56 | 77.08 [71.00, 84.84] | 163/908/58/1013 | 50.00 | 50.00 | 27.08 |
| 1.00 | 100.00±0.00 | 100.00 [100.00, 100.00] | 221/1921/0/0 | 100.00 | 100.00 | 0.00 |

**Table 3:** Comparison of ML risk-ranking vs simple heuristic baselines on GRH TEST set.

| Policy | $K = 0.20$ | $K = 0.30$ | $K = 0.38$ | $K = 0.45$ | $K = 0.50$ |
| --- | --- | --- | --- | --- | --- |
| ML Champion (XGBoost) | £90.94 | £83.24 | £81.42 | £79.08 | £77.08 |
| Shortest lead time | £94.68 | £93.51 | £89.36 | £90.29 | £92.02 |
| Prior canceller $\wedge$ short lead ($\leq 14$ days) | £95.14 | £93.51 | £87.96 | £90.29 | £92.02 |
| Prior canceller $\vee$ short lead ($\leq 14$ days) | £96.08 | £92.58 | £88.42 | £89.36 | £89.68 |
| Random baseline | £103.55 | £102.38 | £100.09 | £100.09 | £99.02 |

Increasing the intervention budget from *K*=0.38 to *K*^⋆^=0.50 raises the intervention cost (more cases reviewed) and reduces missed LCDNA cost, with a reduction in total cost per case on the sealed GRH test set (80.95 [73.15, 90.39] → 77.08 [71.00, 84.84]). The optimal *K*^⋆^=0.50 was selected to minimise DEV mean cost rather than TEST cost, reflecting the sealed test principle.

#### 4.1.1. Cost–Capacity Frontier and Policy Selection

We computed a cost–capacity frontier over a grid of *K* values using DEV folds (CAL+VAL) and selected *K*^⋆^ as the minimiser of the DEV mean cost. The DEV-optimal policy was *K*^⋆^=0.50. For transparency, we note that the minimum observed TEST cost occurred at *K*=0.50 (matching *K*^⋆^); however, policy selection was based on DEV performance only, reflecting the sealed test principle.

### 4.2. Model & Policy Comparison

To assess the relative performance of the champion XGBoost model against interpretable baselines and alternative imbalance-handling strategies, we evaluated six model configurations on the GRH holdout TEST set (sealed) and DEV folds. The models included: (1) Champion XGBoost (hyperparameter-tuned with Optuna), (2) Logistic Regression baseline (with class weighting for interpretability), (3) Random Forest baseline (simple tree ensemble), (4) XGBoost with scale_pos_weight only (no hyperparameter tuning), (5) XGBoost with ADASYN oversampling, and (6) XGBoost with balanced bagging. All models were trained on the same feature set (baseline features only, aligned to common features across all splits) and evaluated at clinically plausible intervention capacities: *K* ∈ {0.20, 0.30, 0.38, 0.45, 0.50}.

Table 4 reports Cost@K on DEV (mean ± std across folds) and TEST (sealed). The champion XGBoost model achieves the lowest TEST cost at *K* = 0.45 (76.75) and *K* = 0.50 (77.08), with competitive performance across all *K* values. Logistic Regression performs surprisingly well, achieving the lowest DEV cost at *K* = 0.38 (76.10±7.06) and *K* = 0.45 (76.32±5.40), but with higher TEST costs (82.35 and 83.75 respectively), indicating potential overfitting. Random Forest shows consistent but slightly higher costs than the champion model. The imbalance-handling variants (scale_pos_weight only, ADASYN, bagging) show mixed results, with scale_pos_weight achieving competitive TEST costs at higher *K* values but ADASYN showing higher variance on DEV.

**Table 4:** Cost@K comparison across models and policies on DEV (mean ± std) and TEST (sealed).

| Model | K=20% |  | K=30% |  | K=38% |  | K=45% |  | K=50% |  |
| --- | --- | --- | --- | --- | --- | --- | --- | --- | --- | --- |
|  | DEV | TEST | DEV | TEST | DEV | TEST | DEV | TEST | DEV | TEST |
| Champion | 85.01 | 90.94 | 78.69 | 85.11 | 78.76 | 80.95 | 77.35 | 76.75 | 76.85 | 77.08 |
| XGB | $\pm 6.63$ | | $\pm 5.41$ | | $\pm 5.90$ | | $\pm 4.33$ | | $\pm 3.56$ | |
| Logistic | 85.25 | 90.01 | 80.76 | 85.57 | 76.10 | 82.35 | 76.32 | 83.75 | 77.08 | 82.21 |
| Reg | $\pm 6.98$ | | $\pm 4.86$ | | $\pm 7.06$ | | $\pm 5.40$ | | $\pm 5.23$ | |
| Random | 85.49 | 91.88 | 81.82 | 87.44 | 78.27 | 82.82 | 78.43 | 82.82 | 79.52 | 81.28 |
| Forest | $\pm 5.64$ | | $\pm 6.07$ | | $\pm 6.93$ | | $\pm 5.21$ | | $\pm 5.07$ | |
| XGB | 86.60 | 90.48 | 82.10 | 84.64 | 79.30 | 83.29 | 76.82 | 81.42 | 77.62 | 83.61 |
| weight | $\pm 8.20$ | | $\pm 6.17$ | | $\pm 6.12$ | | $\pm 5.61$ | | $\pm 4.37$ | |
| XGB | 87.34 | 88.61 | 79.69 | 87.91 | 77.48 | 86.09 | 77.89 | 84.22 | 77.86 | 83.15 |
| ADASYN | $\pm 11.94$ | | $\pm 9.52$ | | $\pm 6.85$ | | $\pm 6.12$ | | $\pm 5.80$ | |
| XGB | 84.42 | 93.74 | 80.43 | 86.97 | 79.29 | 83.29 | 78.65 | 82.35 | 79.79 | 80.81 |
| Bagging | $\pm 3.52$ | | $\pm 6.00$ | | $\pm 4.80$ | | $\pm 6.72$ | | $\pm 4.73$ | |

Leave-one-hospital-out cross-validation provides an auxiliary heterogeneity check on the training hospitals. For the champion XGBoost configuration, LOHO-CV mean cost is £84.51 ± 13.63, with hospital-specific costs of £88.41 at CGH, £62.56 at STG, £100.00 at CIR, and £87.05 at TWC. This spread suggests material site-to-site heterogeneity, with the strongest transfer performance at STG and the weakest at CIR.

Table 5 and Table 6 report Recall@K and Precision@K respectively. Recall (sensitivity) measures the proportion of actual cancellations captured by the intervention policy, while precision measures the proportion of intervened cases that actually cancel. The champion model achieves the highest TEST recall at *K* = 0.50 (0.738), indicating strong ability to identify high-risk cases. However, precision remains modest across all models (0.139–0.173 at *K* = 0.50), reflecting the low base rate of cancellations (approximately 10.3%). Logistic Regression shows competitive recall but lower precision, suggesting it may be over-intervening on lower-risk cases.

**Table 5:**
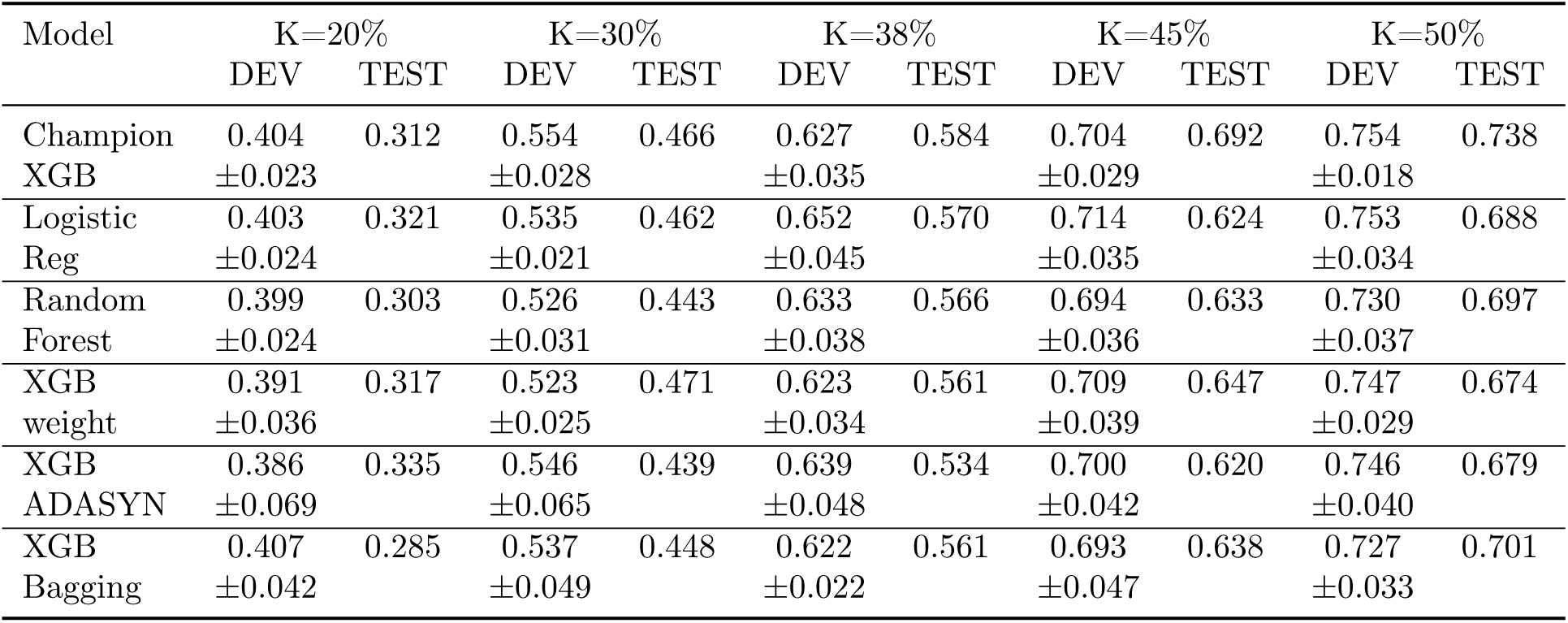
Recall@K comparison across models and policies on DEV (mean ± std) and TEST (sealed).

**Table 6:** Precision@K comparison across models and policies on DEV (mean ± std) and TEST (sealed).

| Model | K=20% |  | K=30% |  | K=38% |  | K=45% |  | K=50% |  |
| --- | --- | --- | --- | --- | --- | --- | --- | --- | --- | --- |
|  | DEV | TEST | DEV | TEST | DEV | TEST | DEV | TEST | DEV | TEST |
| Champion | 0.220 | 0.161 | 0.201 | 0.160 | 0.180 | 0.158 | 0.170 | 0.159 | 0.164 | 0.152 |
| XGB | $\pm 0.013$ | | $\pm 0.014$ | | $\pm 0.011$ | | $\pm 0.013$ | | $\pm 0.010$ | |
| Logistic | 0.219 | 0.166 | 0.194 | 0.159 | 0.186 | 0.155 | 0.173 | 0.143 | 0.164 | 0.142 |
| Reg | $\pm 0.010$ | | $\pm 0.014$ | | $\pm 0.012$ | | $\pm 0.011$ | | $\pm 0.009$ | |
| Random | 0.217 | 0.157 | 0.190 | 0.152 | 0.181 | 0.154 | 0.168 | 0.145 | 0.159 | 0.144 |
| Forest | $\pm 0.021$ | | $\pm 0.013$ | | $\pm 0.005$ | | $\pm 0.012$ | | $\pm 0.012$ | |
| XGB | 0.212 | 0.164 | 0.190 | 0.162 | 0.178 | 0.152 | 0.171 | 0.148 | 0.163 | 0.139 |
| weight | $\pm 0.012$ | | $\pm 0.008$ | | $\pm 0.010$ | | $\pm 0.012$ | | $\pm 0.011$ | |
| XGB | 0.208 | 0.173 | 0.198 | 0.151 | 0.183 | 0.145 | 0.169 | 0.142 | 0.162 | 0.140 |
| ADASYN | $\pm 0.027$ | | $\pm 0.020$ | | $\pm 0.015$ | | $\pm 0.011$ | | $\pm 0.010$ | |
| XGB | 0.223 | 0.147 | 0.195 | 0.154 | 0.178 | 0.152 | 0.167 | 0.146 | 0.158 | 0.145 |
| Bagging | $\pm 0.037$ | | $\pm 0.025$ | | $\pm 0.011$ | | $\pm 0.012$ | | $\pm 0.011$ | |

Overall, the champion XGBoost model demonstrates superior performance in the cost-sensitive setting, with the lowest TEST costs at *K* = 0.45 (76.75) and *K* = 0.50 (77.08) and strong recall-precision balance. Logistic Regression provides a competitive interpretable alternative, achieving the lowest DEV costs at *K* = 0.38 and *K* = 0.45 but with higher TEST costs, suggesting some overfitting. Random Forest shows consistent performance but slightly higher costs than the champion. The results suggest that hyperparameter tuning and careful imbalance handling (via scale_pos_weight) are more effective than oversampling or bagging strategies for this task.

#### 4.2.1. Comparison with Simple Heuristic Baselines

To contextualise the value of ML-based risk ranking, we compared the champion XGBoost model against simple, operationally transparent heuristic baselines evaluated on the same *K* grid. The heuristics tested were: (1) *shortest lead time*: intervene on cases with the shortest booking-to-procedure lead time; (2) *prior canceller* ∧ *short lead* : intervene on cases that are prior cancellers *and* have short lead time (≤14 days); (3) *prior canceller* ∨ *short lead* : intervene on cases that are prior cancellers *or* have short lead time (≤14 days); and (4) *random baseline*: random selection (for comparison). All heuristics were evaluated on the sealed GRH TEST set using the same top-*K* policy framework and cost model.

Table 3 reports Cost@K for the ML champion and all heuristic baselines. The ML model consistently outperforms all heuristics across all *K* values, with cost savings ranging from 3.9% (at *K* = 0.20) to 14.1% (at *K* = 0.50) compared to the best-performing heuristic. At the optimal policy (*K*^⋆^ = 0.50), the ML model achieves a cost of £77.08 per case, compared to £89.68 for the best heuristic (prior canceller ∨ short lead), representing a savings of £12.60 per case (14.1% reduction). The shortest lead-time heuristic, which is operationally intuitive and commonly used in practice, achieves costs of £89.36–£94.68 across the *K* range, consistently higher than the ML model.

These results demonstrate that ML-based risk ranking provides substantial value beyond simple rule-based approaches. The heuristics capture intuitive risk factors (short lead time, prior cancellation history) but fail to leverage the complex interactions and additional signals (e.g., specialty effects, calendar patterns, patient history) that the ML model learns. The consistent performance gap across all *K* values suggests that the ML advantage is robust to different intervention capacity constraints, making it operationally valuable even when intervention resources are limited.

### 4.3. Secondary Evaluation: Time-Forward (Deployment-Like) Split

In the time-forward evaluation, patients are split chronologically by earliest procedure date into TRAIN / CAL / VAL / TEST. Under the default 60 / 15 / 15 / 10 split, the DEV-optimal budget was *K*^⋆^=0.38, and the sealed TEST cost was £70.97 [63.62, 80.26], substantially lower than in the hospital-holdout setting (77.08 [71.00, 84.84]), indicating that temporal generalisation within the same service environment is easier than cross-hospital domain shift.

Across these plausible split configurations, TEST costs varied within a narrow range (70.97 to 74.02; ≈4.3% variation), supporting the robustness of the time-forward conclusions.

#### 4.3.1. Variable Importance and SHAP-based Explainability

To understand which features drive predictions in both the hospital-holdout (domain-shift) and time-forward (temporal) evaluation settings, we computed SHAP (SHapley Additive exPlanations) values for the champion XGBoost model in both scenarios. Table 8 reports the top 15 features ranked by mean absolute SHAP importance for each model. The feature rankings show strong agreement between the two evaluation settings (rank correlation: 0.886, SHAP value correlation: 0.939), indicating that the model relies on similar predictive signals regardless of whether it is evaluated under domain shift or temporal generalisation.

**Table 7:** Time-forward sensitivity analysis across three scenarios.

| Scenario | Ratios<br>T/C/V/Te | $K^*$ | DEV<br>mean±std | TEST<br>cost | TP/FP/FN/TN | TEST<br>range |
| --- | --- | --- | --- | --- | --- | --- |
| S1 | 0.60/0.15/0.15/0.10 | 0.38 | 75.91±6.16 | 70.97 | 107/574/59/1051 | 03-18<br>04-29 |
| S2 | 0.70/0.10/0.10/0.10 | 0.55 | 78.06±7.14 | 74.02 | 132/853/34/771 | 03-18<br>04-29 |
| S3 | 0.60/0.10/0.10/0.20 | 0.50 | 76.73±3.49 | 71.97 | 262/1560/80/1741 | 02-10<br>04-29 |

**Table 8:** Top 15 features by SHAP importance: Hospital-Holdout vs Time-Forward models.

| Feature | HH Rank | TF Rank | HH SHAP | TF SHAP |
| --- | --- | --- | --- | --- |
| Days between booking and procedure | 1 | 1 | 1.000 | 1.000 |
| procedure_month | 2 | 2 | 0.512 | 0.386 |
| IS_cancer_binary | 4 | 4 | 0.393 | 0.282 |
| Days To Public Holiday | 6 | 3 | 0.357 | 0.299 |
| speciality_Upper_Gastrointestinal_Surgery | 3 | 10 | 0.435 | 0.186 |
| procedure_day | 8 | 7 | 0.241 | 0.209 |
| Days From Public Holiday | 7 | 9 | 0.311 | 0.198 |
| Monday | 5 | 11 | 0.360 | 0.147 |
| speciality_Urology | 10 | 6 | 0.210 | 0.249 |
| patient_cancellation_rate | 9 | 8 | 0.229 | 0.209 |
| Saturday | 16 | 5 | 0.144 | 0.262 |
| speciality_Trauma_and_Orthopaedics | 11 | 13 | 0.185 | 0.132 |
| speciality_Gynaecology | 14 | 14 | 0.161 | 0.131 |
| Days_booking_procedure_bucket | 17 | 12 | 0.120 | 0.134 |
| speciality_Breast_Surgery | 13 | 16 | 0.164 | 0.109 |

The most important feature in both models is *Days between booking and procedure* (lead time), which aligns with operational intuition that shorter lead times are associated with higher cancellation risk. Other consistently important features include *procedure_month*, *IS_cancer_binary*, and temporal features related to public holidays. Notable differences include *Saturday* (rank 16 in hospital-holdout vs rank 5 in time-forward) and *speciality_Upper_Gastrointestinal_Surgery* (rank 3 in hospital-holdout vs rank 10 in time-forward), suggesting that some features may be more domain-specific while others are more temporally stable.

To move beyond a qualitative “feature importance” statement, we quantified the marginal relationship between booking-to-procedure lead time and LCDNA risk using bootstrap estimates for both the hospital-holdout and time-forward models. Table 9 and Figure 4 show the estimated LCDNA risk by lead-time bucket with bootstrap 95% confidence intervals. Both models show a general trend of decreasing risk with increasing lead time, though the patterns differ slightly between evaluation settings. In the hospital-holdout model, risk is highest at 7 days (21.1%) and decreases to 8.0% for 30+ days. In the time-forward model, risk peaks at 8–14 days (15.6%) and decreases to 6.2% for 30+ days. The steepest risk increase occurs when lead time compresses into the 7–14 day range, supporting a clear operational interpretation: cases scheduled within the next two weeks represent a narrow, high-leverage window for targeted confirmation and rescheduling support.

**Table 9:** Estimated LCDNA risk by lead-time bucket (bootstrap 95% CIs): Hospital-Holdout vs Time-Forward models.

| Lead-time (days) | Hospital-Holdout |  | Time-Forward |  |
| --- | --- | --- | --- | --- |
|  | Risk | 95% CI | Risk | 95% CI |
| 7 | 21.1% | [14.0, 28.9] | 12.1% | [5.1, 20.7] |
| 8–14 | 11.5% | [8.5, 14.4] | 15.6% | [11.7, 19.8] |
| 15–30 | 10.3% | [8.4, 12.3] | 9.3% | [7.4, 11.5] |
| 30+ | 8.0% | [6.1, 10.1] | 6.2% | [4.5, 7.9] |

This shift and difference between two models is expected because the two evaluation designs stress different forms of generalisation. Hospital-holdout reflects site/domain differences (e.g., scheduling practices and specialty mix), while time-forward reflects temporal changes in booking behaviour and service configuration. The time-forward test period exhibits a different lead-time distribution and case-mix, which shifts where cancellations concentrate along the lead-time axis. Despite this difference in the modal peak, both evaluations consistently identify the 7–14 day interval as the most operationally actionable window for targeted outreach, suggesting that hospitals should calibrate outreach timing to their local lead-time profile and monitor drift over time.

To support clinical interpretability and to validate that the learned signals align with operational intuition, we analysed model explainability using three complementary global importance views: (i) XGBoost internal gain-based feature importance, (ii) permutation importance under the operational policy objective (cost@K, with *K*^⋆^=0.50 for hospital-holdout and *K*^⋆^=0.38 for time-forward), and (iii) SHAP values for the final XGBoost champion model evaluated in both hospital-holdout and time-forward settings. Table 10 reports the top five specialties by SHAP importance for both models, along with observed LCDNA rates and median lead times from the GRH validation split.^1^

**Table 10:** Top specialties by mean absolute SHAP contribution on development data, with observed LCDNA rate and median lead time: Hospital-Holdout vs Time-Forward models.

| Specialty | SHAP |  | N | Rate | 95% CI | Med LT |
| --- | --- | --- | --- | --- | --- | --- |
|  | HH | TF |  |  |  |  |
| Upper_Gastrointestinal_Surgery | 0.134 | 0.088 | 238 | 4.6% | [2.1, 7.6] | 28 |
| Urology | 0.111 | 0.089 | 79 | 10.1% | [3.8, 17.7] | 21 |
| Breast_Surgery | 0.071 | 0.088 | 75 | 1.3% | [0.0, 4.0] | 17 |
| Trauma_and_Orthopaedics | 0.075 | 0.082 | 93 | 21.5% | [14.0, 30.1] | 28 |
| ENT | 0.098 | 0.054 | 666 | 12.8% | [10.5, 15.5] | 17 |

To further understand how temporal and calendar features contribute to predictions across evaluation settings, we examined date-related features using SHAP analysis for both models. Table 11 reports the SHAP importance of date-related features, categorised by type (month, day of week, holiday proximity, etc.). The most important date feature in both models is *Days between booking and procedure* (lead time), with SHAP values of 0.420 (hospital-holdout) and 0.403 (time-forward), confirming its dominant role in cancellation risk prediction.

**Table 11:** Date-related features by mean absolute SHAP contribution: Hospital-Holdout vs Time-Forward models.

| Feature | SHAP |  | Category |
| --- | --- | --- | --- |
|  | HH | TF |  |
| Days between booking and procedure | 0.420 | 0.403 | Lead Time |
| procedure_month | 0.082 | 0.313 | Month |
| procedure_day | 0.107 | 0.126 | Day of Month |
| Days From Public Holiday | 0.164 | 0.059 | Holiday Proximity |
| Monday | 0.129 | 0.093 | Day of Week |
| Days From School Holiday | 0.127 | 0.065 | Holiday Proximity |
| Days_booking_procedure_bucket | 0.087 | 0.082 | Lead Time |
| Days To School Holiday | 0.084 | 0.084 | Holiday Proximity |
| Saturday | 0.045 | 0.107 | Day of Week |
| Days To Public Holiday | 0.077 | 0.048 | Holiday Proximity |

Notable differences emerge between the two evaluation settings. *procedure_month* shows substantially higher importance in the time-forward model (0.313) compared to hospital-holdout (0.082), suggesting that seasonal patterns are more predictive when evaluating temporal generalisation within the same service environment. Conversely, *Days From Public Holiday*is more important in the hospital-holdout setting (0.164 vs 0.059), potentially reflecting domain-specific operational patterns. Day-of-week effects also differ: *Saturday* is more important in time-forward (0.107 vs 0.045), while *Monday*shows higher importance in hospital-holdout (0.129 vs 0.093). These differences highlight that while lead time remains the primary temporal signal, the relative importance of calendar effects varies between domain-shift and temporal evaluation settings.

Finally, to summarise interpretability at a higher level, we grouped features into clinically meaningful categories and observed that *lead time* and *specialty* dominate total attribution mass, followed by comorbidity burden, day-of-week, and patient-history features. This grouped view is helpful for communicating model behaviour to clinical stakeholders without over-emphasising individual one-hot indicators. Figure 5 presents SHAP summary (beeswarm) plots computed on stratified validation subsets, where each point corresponds to a case–feature attribution and colour encodes the underlying feature value. Positive SHAP values increase the predicted LCDNA risk, while negative values decrease it. Across both the hospital-holdout and time-forward settings, the beeswarm patterns indicate that risk ranking is driven by a small set of recurring, manager- and clinician-interpretable factors rather than a single variable; at the same time, modest shifts in the spread and ordering of attributions between the two panels are consistent with the different stresses induced by site/domain shift versus temporal shift.

We further summarise global importance using mean absolute SHAP values (bar plots), which capture each feature’s average contribution magnitude while leaving directionality to the beeswarm plots. Because probability-based feature importance does not necessarily reflect sensitivity of the *deployment policy*, we additionally compute permutation importance with respect to the operational objective (cost@K) (Figure 7), at the selected operating points (*K*^⋆^ = 0.50 for hospital-holdout and *K*^⋆^ = 0.38 for time-forward). Features whose permutation causes the largest increase in cost@K are those to which the intervention policy is most sensitive, providing a decision-centric audit that complements SHAP. To characterise functional relationships for the dominant drivers, Figure 8 shows SHAP dependence plots for the top three features (ranked by mean absolute SHAP), highlighting potentially non-linear and threshold-like effects that help explain why multivariate ranking can outperform simple heuristics. Finally, to reduce fragmentation introduced by one-hot encoding and to improve interpretability for clinical stakeholders, we aggregate SHAP importance into clinically meaningful feature groups (Figure 9; e.g., lead-time, specialty, day-of-week, patient-history, comorbidity), enabling a clear communication of the principal factors underpinning model decisions while preserving quantitative attribution structure.

**Figure 6:**
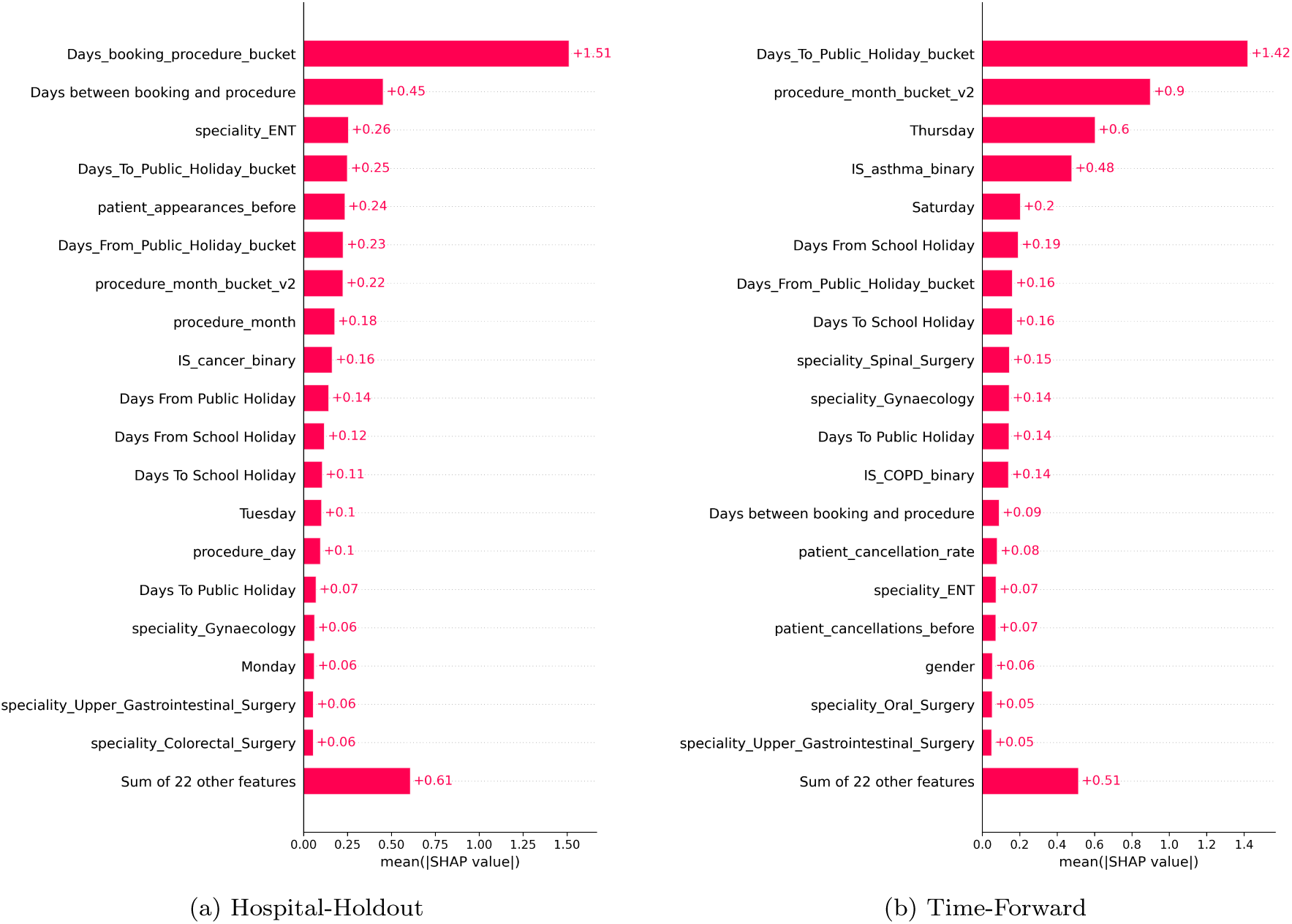
Mean absolute SHAP values (feature importance)

**Figure 7:**
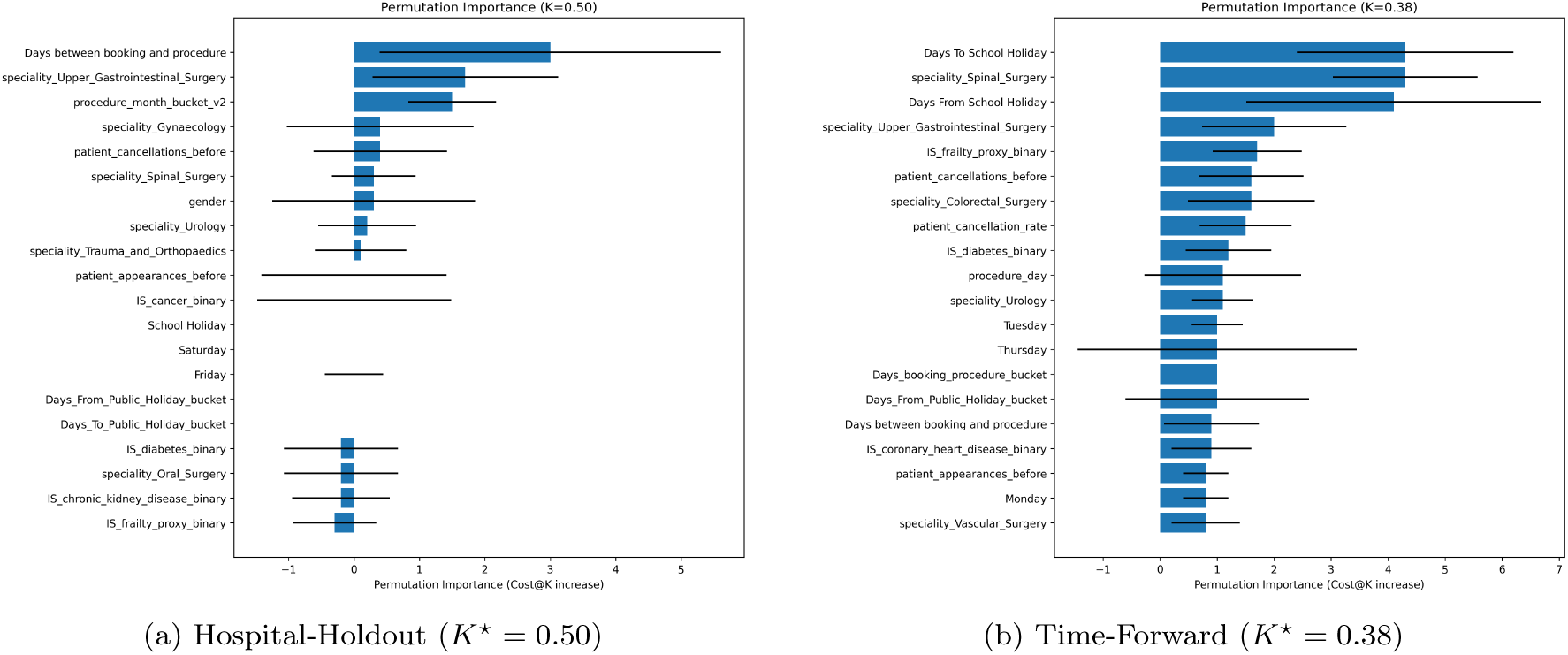
Permutation importance under the cost-sensitive objective

**Figure 8:**
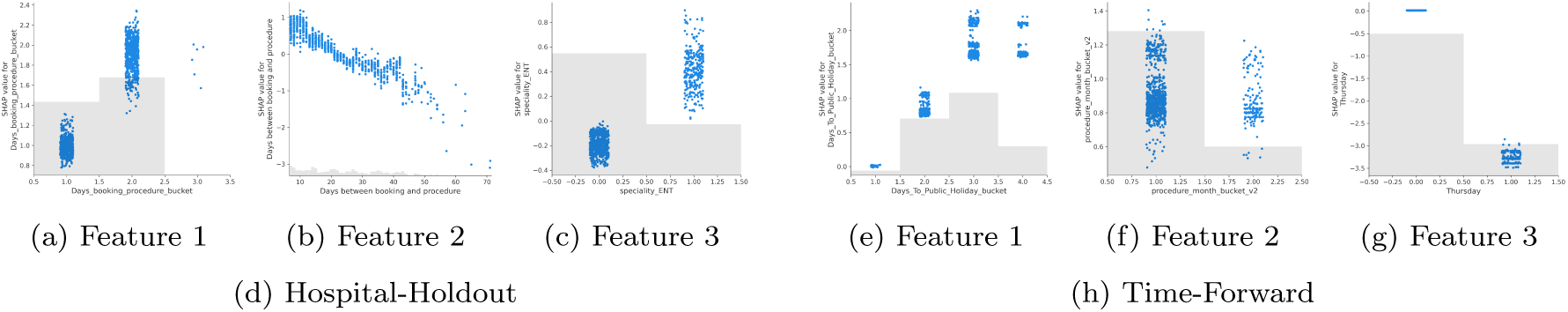
SHAP dependence plots for the top three most important features

**Figure 9:**
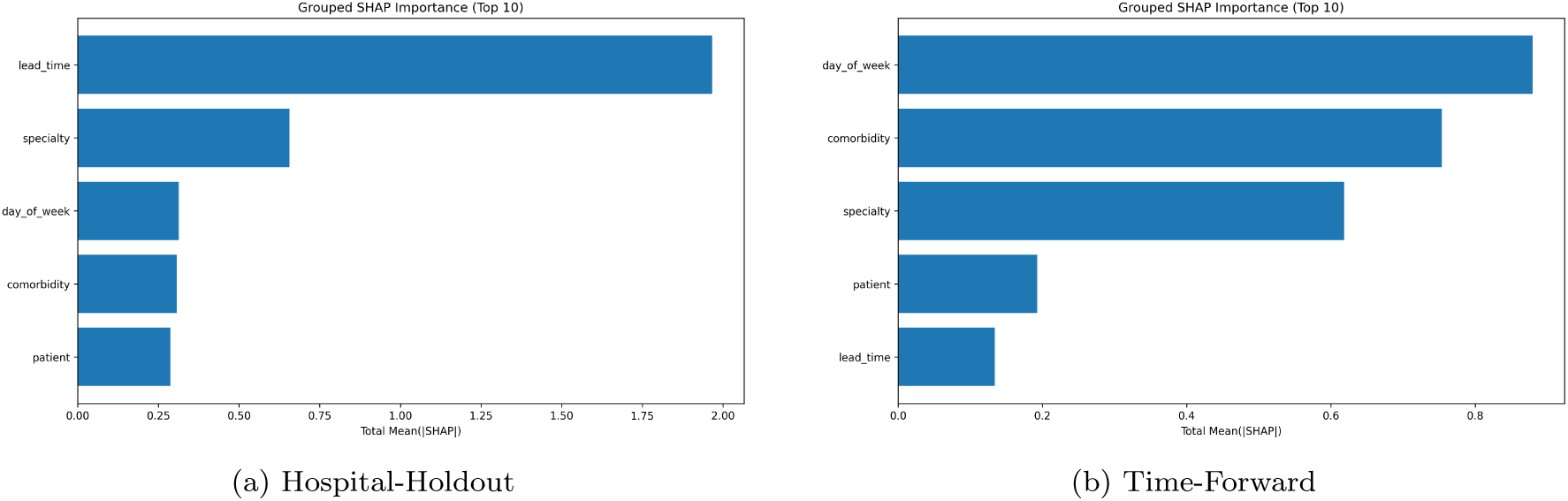
Grouped SHAP importance by feature category

#### 4.3.2. Comprehensive Sensitivity Analysis: Cost Ratio and Intervention Effectiveness

Our cost model makes two key assumptions: (i) a cost ratio *m/i* = 10 (where *m* = *£*1,000 and *i* = *£*100), and (ii) perfect intervention effectiveness (*e* = 1.0), meaning that true positives avoid the full avoidable cost *m* with no residual cost. To assess the robustness of our conclusions, we conducted a comprehensive sensitivity analysis varying both the cost ratio *m/i* ∈ {5, 7.5, 10, 12.5, 15} and intervention effectiveness *e* ∈ {0.50, 0.75, 1.0}.

Under partial effectiveness, the modified cost function becomes:

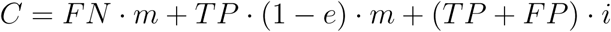

where *e* = 1.0 represents perfect effectiveness (current assumption) and *e* = 0.0 represents no effectiveness.

Figure 10 shows six panels summarising the sensitivity analysis: (a) optimal *K*^∗^ vs cost ratio for different effectiveness levels, (b) optimal cost vs cost ratio, (c) cost savings percentage vs cost ratio, (d) optimal *K*^∗^ vs effectiveness for different cost ratios, (e) optimal cost vs effectiveness, and (f) a heatmap showing *K*^∗^ stability across the parameter space. Table 12 reports key results for selected scenarios.

**Figure 10:**
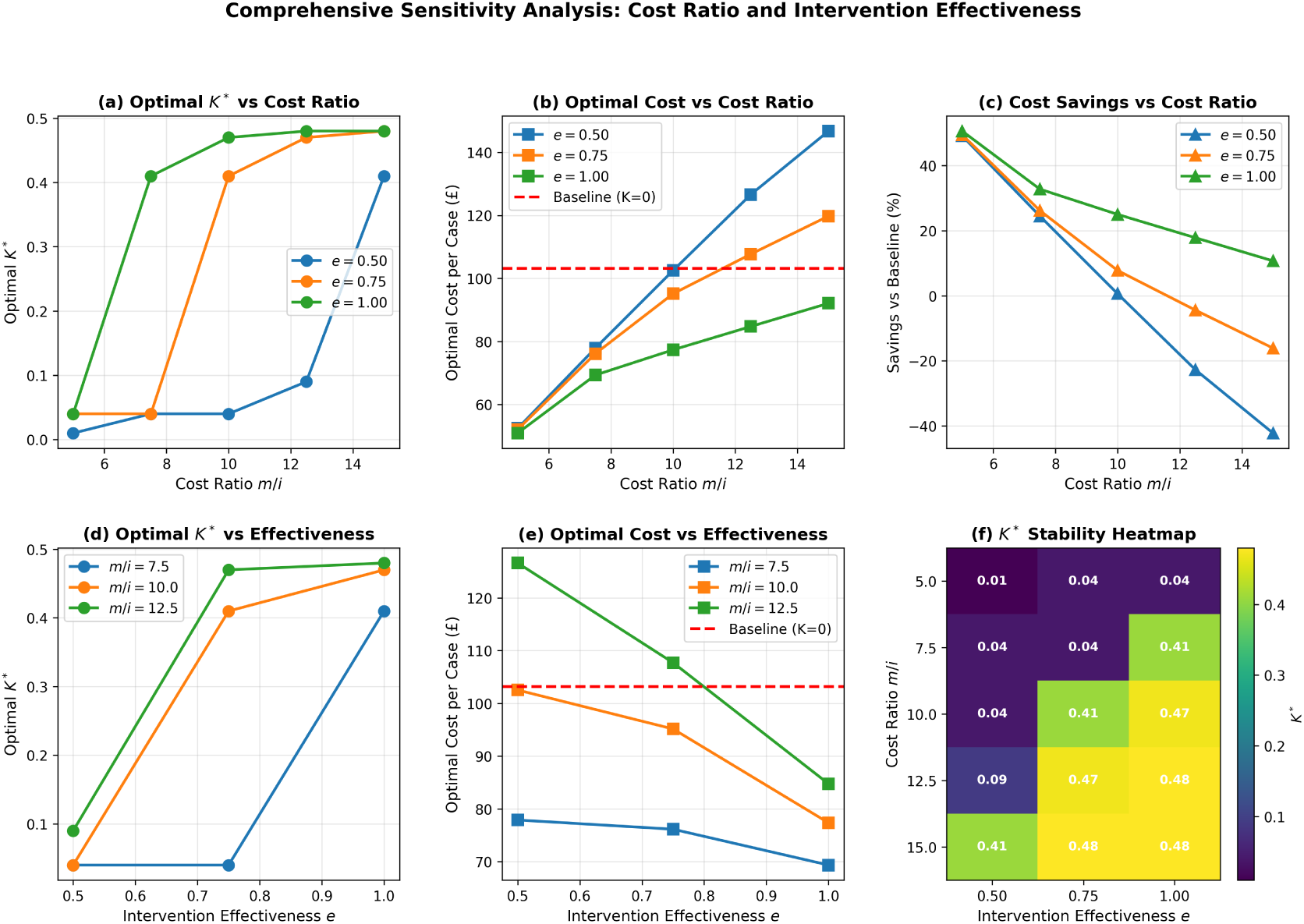
Comprehensive sensitivity analysis: (a) optimal *K*^∗^ vs cost ratio, (b) optimal cost vs cost ratio, (c) savings percentage vs cost ratio, (d) optimal *K*^∗^ vs effectiveness, (e) optimal cost vs effectiveness, (f) *K*^∗^ stability heatmap. The baseline do-nothing cost (£103.17 per case) is shown as a dashed red line in panels (b) and (e).

**Table 12:** Sensitivity analysis: optimal *K*^∗^ and cost savings across cost ratios and intervention effectiveness.

| $m/i$ | $e$ | $K^*$ | Optimal Cost<br>(£/case) | Savings<br>(£/case) | Savings<br>(%) |
| --- | --- | --- | --- | --- | --- |
| 7.5 | 0.75 | 0.04 | 76.14 | 27.03 | 26.2 |
| 7.5 | 1.00 | 0.41 | 69.35 | 33.82 | 32.8 |
| 10.0 | 0.75 | 0.41 | 95.14 | 8.03 | 7.8 |
| 10.0 | 1.00 | 0.47 | 77.36 | 25.82 | 25.0 |
| 12.5 | 0.75 | 0.47 | 107.70 | -4.53 | -4.4 |
| 12.5 | 1.00 | 0.48 | 84.76 | 18.42 | 17.9 |

The analysis demonstrates three key findings. First, *optimal K*^∗^ *is relatively stable* across plausible cost ratios: for *e* = 1.0, *K*^∗^ ranges from 0.41 to 0.48 as *m/i* varies from 7.5 to 12.5, with the base case (*m/i* = 10, *e* = 1.0) yielding *K*^∗^ = 0.47. Second, *cost savings remain positive* across a wide range of assumptions: for *m/i* ≥ 7.5 and *e* ≥ 0.75, the optimal policy achieves cost savings of 7.8%–32.8% compared to the do-nothing baseline (£103.17 per case). Third, *the qualitative conclusion that targeted intervention reduces cost* holds even under conservative assumptions: at *m/i* = 7.5 and *e* = 0.75, the optimal policy still achieves 26.2% savings (£27.03 per case).

The sensitivity analysis also reveals that lower effectiveness (*e <* 0.75) or lower cost ratios (*m/i <* 7.5) shift the optimal policy toward lower intervention capacity (*K*^∗^ *<* 0.10), reflecting the reduced economic value of interventions under these assumptions. However, for the base case assumptions (*m/i* = 10, *e* = 1.0), the optimal policy at *K*^∗^ = 0.47 is robust and aligns closely with the development-selected recommendation (*K*^⋆^ = 0.50).

**Operational interpretation of** *K*^⋆^ = 0.50. The optimal policy (*K*^⋆^ = 0.50) means intervening on the top 50% of cases by predicted risk. For a typical medium-sized NHS hospital performing approximately 50 elective procedures per week, this translates to approximately 25 interventions per week (5 per working day). Assuming each intervention takes 10–15 minutes (telephone/virtual contact plus reminders), this requires approximately 5–6 hours of staff time per week, or roughly 1 hour per day. This workload is operationally feasible for a dedicated pre-operative coordination team and represents a manageable intervention capacity that can be scaled up or down based on available resources and seasonal demand.

#### 4.3.3. Variable Importance in Comparison

To improve interpretability and support operational trust, we analysed feature importance using complementary views: (i) native XGBoost split-gain importance, (ii) permutation importance, and (iii) SHAP (TreeSHAP) explanations computed on held-out evaluation samples. Across analyses, the most influential predictors consistently reflected clinically and operationally plausible signals, including lead-time (days between booking and procedure), patient history (e.g., prior cancellation behaviour / cancellation rate), specialty indicators, calendar effects (day-of-week and holiday proximity), and key comorbidity flags such as cancer status. These patterns are consistent with the top-10 lists extracted for gain / permutation / SHAP on the GRH validation sample (see Figure 11)

**Figure 11:**
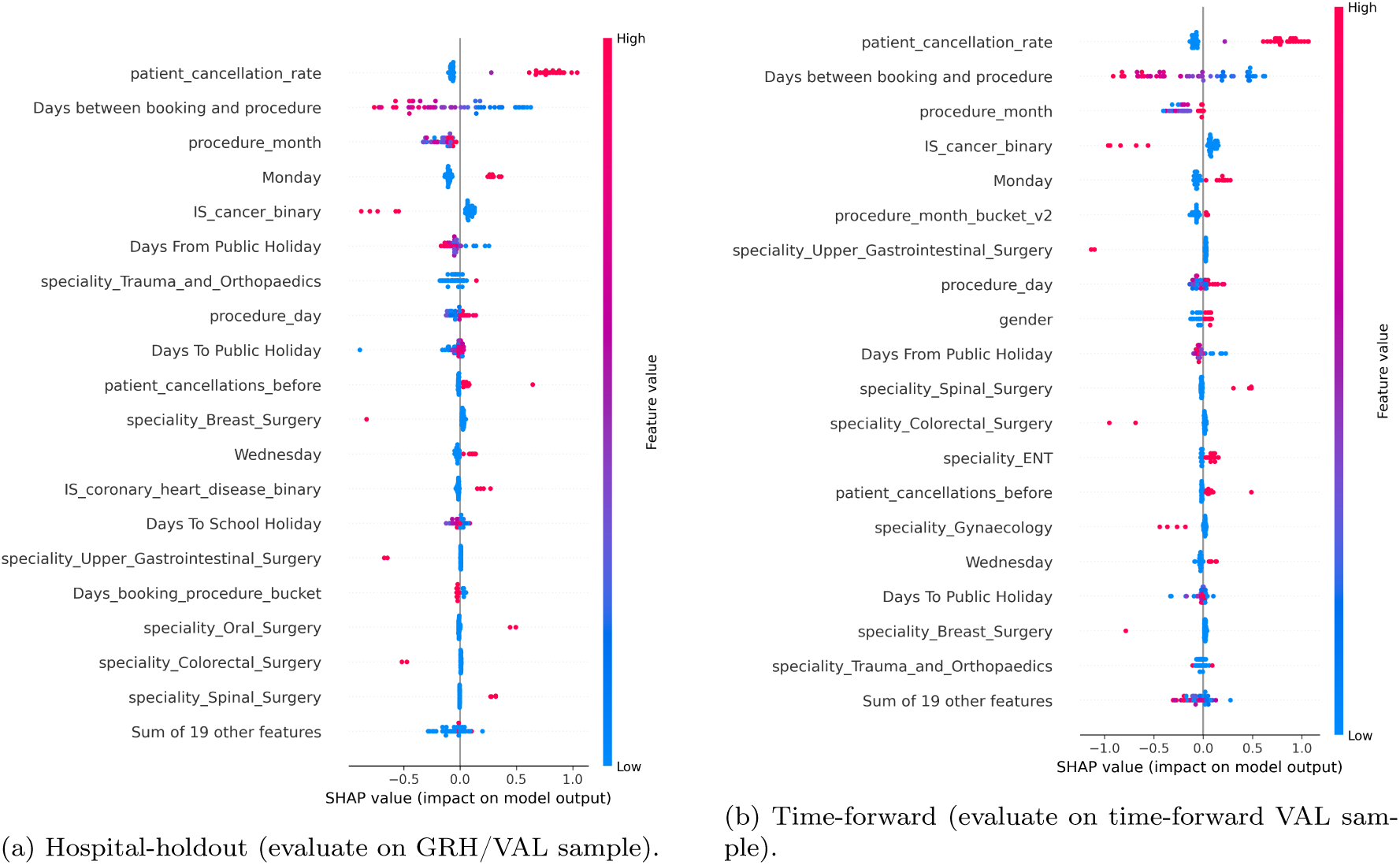
SHAP summary plots (TreeSHAP) for the champion XGBoost model under two evaluation regimes. Features are ordered by mean absolute SHAP value; points show the distribution of per-case contributions.

**Figure 12:**
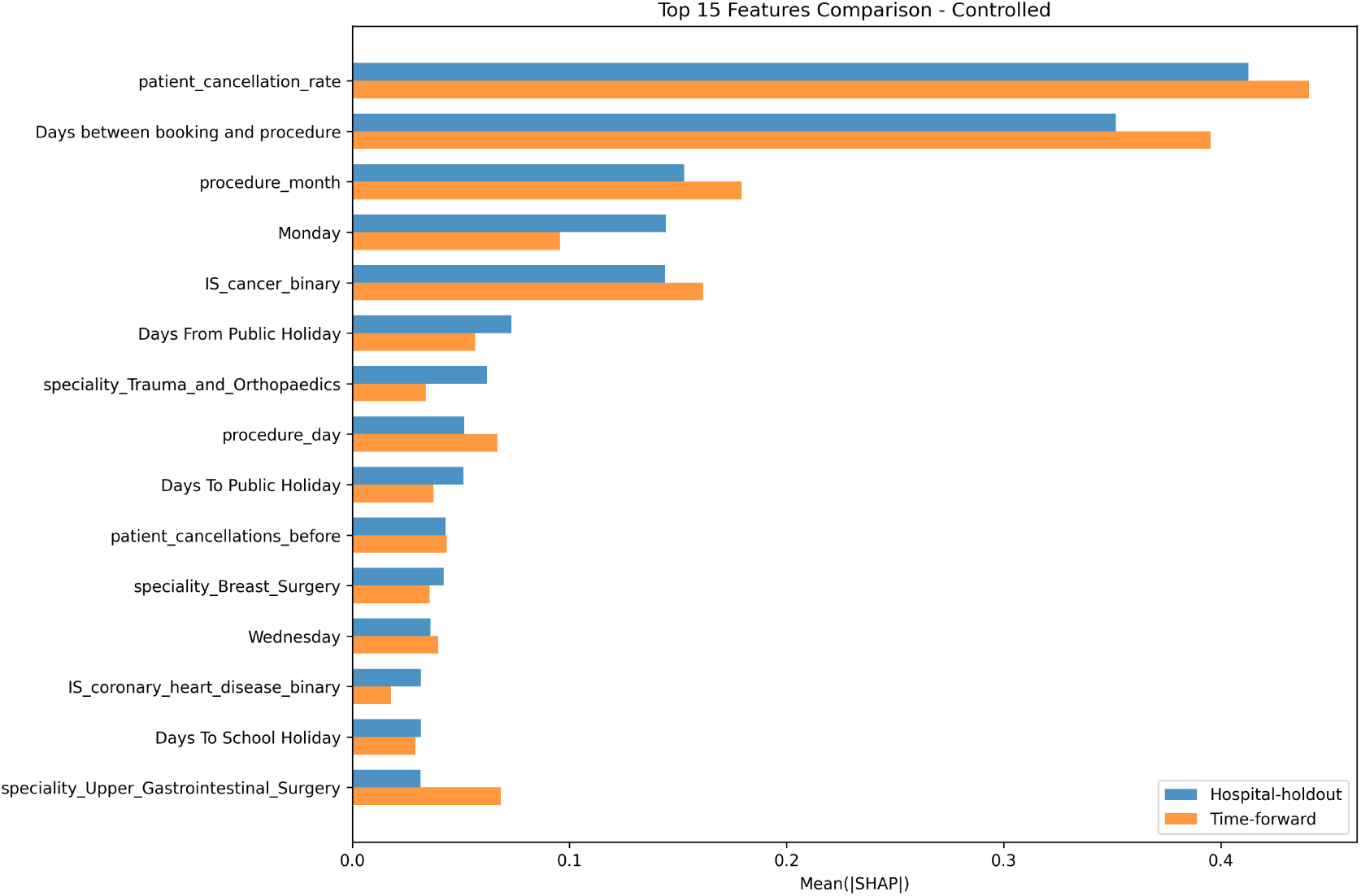
Comparison of feature-importance ranks between hospital-holdout and time-forward regimes. The plot highlights features with the largest rank changes (delta-rank), indicating sensitivity to spatial vs temporal generalisation.

**Figure 13:**
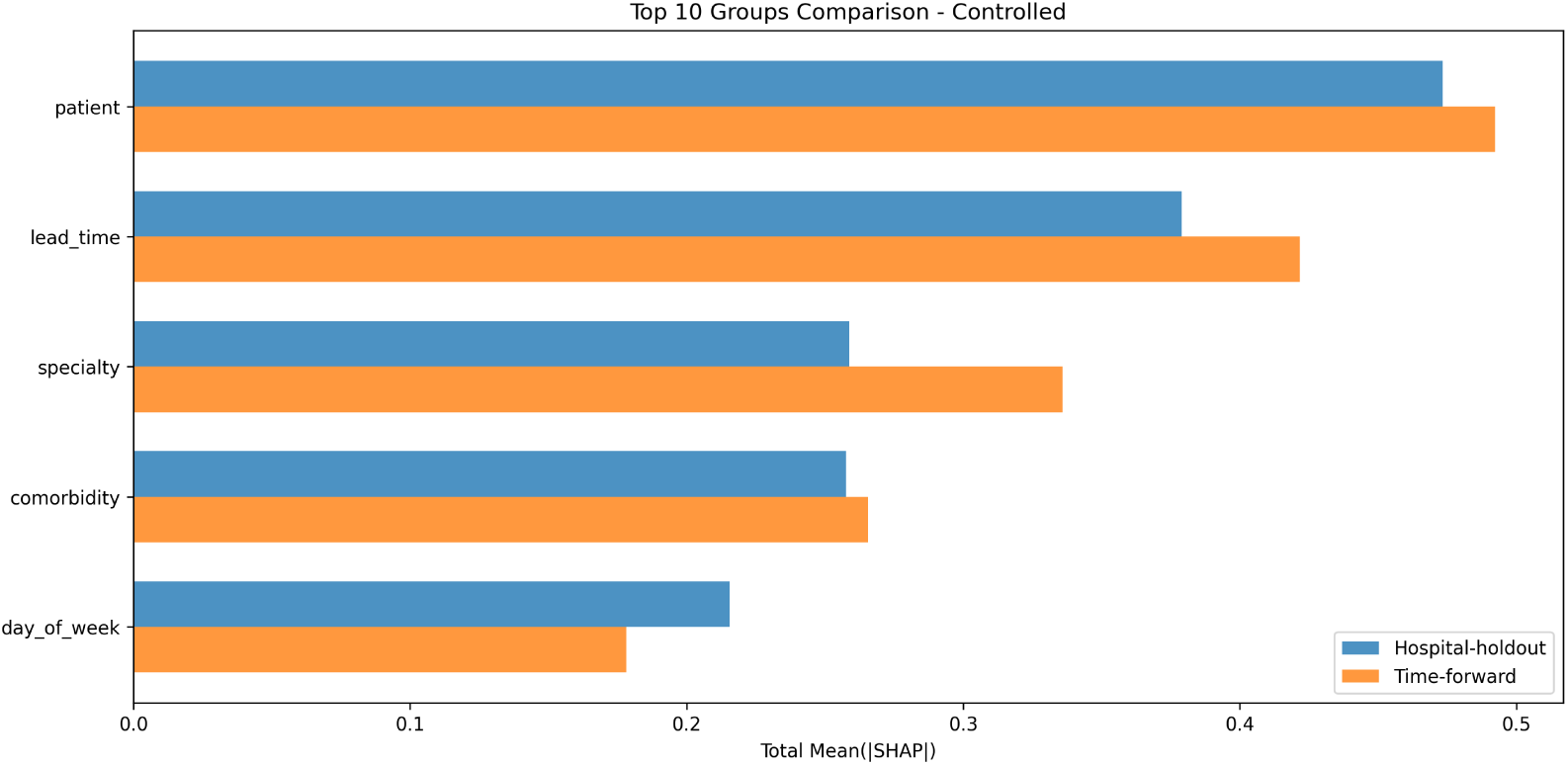
group comparison controlled

##### Hospital-holdout vs time-forward importance

We next compared explainability under two evaluation regimes: *hospital-holdout* (training on non-holdout hospitals, evaluating on GRH) and *time-forward* (training on earlier bookings, evaluating on later bookings). The top features remained broadly consistent between regimes (e.g., lead-time, specialty mix, and patient-history signals), but notable rank shifts were observed. In the contextual comparison, hospital-holdout explanations placed greater weight on specialty composition (e.g., Breast/Oral/Colorectal surgery indicators), whereas time-forward explanations elevated calendar/seasonality-related features (e.g., procedure month, school-holiday proximity) and demographic factors (e.g., gender) into the top ranks. These differences likely reflect the dominant generalisation challenge: spatial case-mix variation across hospitals versus temporal covariate shift across booking periods.

##### Controlled comparison to reduce sampling artefacts

To partially decouple regime effects from sampling differences, we additionally computed a controlled comparison on a small matched set of rows (shared across both regimes). In this controlled set, patient-history and lead-time remained dominant, while several features exhibited substantial rank movement (e.g., gender, procedure-month bucketisation, and specific specialties such as ENT / Gynaecology / Upper GI / Spinal surgery). Given the controlled set is small (N=52), these rank changes should be interpreted cautiously and treated as indicative rather than definitive; nevertheless, they align with the intuition that time-forward evaluation is more sensitive to calendar structure, while hospital-holdout evaluation is more sensitive to cross-site differences in specialty mix and pathways.

## 5. Discussion

### 5.1. Principal findings

This study develops and audits a cost-sensitive machine learning pipeline for predicting late cancellations and no-shows (LCDNA) in elective surgery, with evaluation designed to reflect deployment constraints in NHS theatres. Rather than focusing on discrimination metrics alone, we treat the problem as an operational policy question in which missed LCDNA events carry a high cost and proactive interventions carry a lower one. The framework combines an XGBoost risk model with a capacity-aware top-*K* intervention policy selected on development data only.

The core empirical finding is that targeted intervention remains economically preferable to both doing nothing and intervening on all cases, and that this conclusion holds under both cross-site and time-forward evaluation. The hospital-holdout regime provides the more conservative estimate of deployment performance, whereas the time-forward regime suggests better performance when the care environment is stable. Together, these results support two practical conclusions: first, risk-based ranking is operationally useful; and second, generalisation across hospitals is more challenging than generalisation forward in time within the same service context.

### 5.2. Why domain shift matters for deployment

A key practical contribution of this work is the explicit evaluation of two distinct generalisation challenges: (i) *spatial* shift across hospital sites (case-mix, specialty composition, operational pathways, and local processes), and (ii) *temporal* shift as booking patterns evolve over time. The observed cost gap between hospital-holdout and time-forward evaluation reinforces why random splits can be optimistic for operational modelling. By enforcing patient-level exclusivity and using a sealed test principle, the protocols used here provide a conservative estimate of expected deployment performance when models are applied to new sites or new time windows.

From an implementation perspective, the hospital-holdout results suggest that a Trust-wide model is best treated as an initial baseline that may require local adaptation. In practice, hospitals can update the policy parameter *K* (and, if necessary, recalibrate or refit the model) using recent local development data without ever using future outcomes from the intended deployment window. This approach aligns with the evaluation workflow and reduces the risk of overfitting policy decisions to a single retrospective test set.

### 5.3. Deployment guidance: domain-shift risk budget and phased roll-out

The difference between the hospital-holdout and time-forward cost estimates (£77.08 vs £70.97) quantifies the additional uncertainty introduced when moving across hospital sites. For Trust-wide deployment, we recommend treating this £6.11 per-case gap as a domain-shift risk budget, i.e., the minimum savings that should be promised at go-live for a new site before local optimisation.

A phased roll-out can protect these guaranteed savings while creating a clear route to improvement: (1) start each new site with a conservative Top-*K* aligned to local capacity and monitor Cost@K, missed-LCDNA counts, and workload volume weekly; (2) recalibrate *K* and/or cost parameters during an initial stabilisation period (e.g., 8–12 weeks) using local operational feedback; and (3) only then pursue additional gains toward the within-site benchmark (≈£70.97 per case) through specialty-specific workflow redesign, earlier confirmation, and pathway fixes.

Framing the cross-site result as the “guaranteed” baseline avoids over-promising, while the time-forward benchmark becomes an optimisation target that implementation teams can work toward once local processes are understood.

### 5.4. Operational implications of the Top-K policy

Our aim is not prediction for its own sake but the allocation of limited “high-touch” interventions (e.g., telephone confirmation, transport/problem-solving support, pre-operative checks, and rapid rescheduling) to the cases where they yield the greatest expected operational benefit. We therefore operationalise risk scores through a top-*K* policy: for each planning horizon, rank upcoming cases by estimated LCDNA risk/expected cost and intervene on the highest-risk *K* cases, subject to capacity.

Selecting *K* makes workload explicit and allows decision makers to trade off savings against staff time. *K* should be set locally based on available staffing, lead-time constraints, and service-line workflow, and then reviewed routinely as baseline cancellation rates and operational conditions change. In practice this means monitoring Cost@K alongside missed-LCDNA counts (false negatives) and weekly contact volumes, and adjusting *K* when constraints tighten or when pathway changes reduce avoidable cancellations.

### 5.5. Tiered intervention workflows and pathway audit

To translate model outputs into action, we recommend tiered intervention workflows that are driven by both risk ranking and operational urgency. Tier 1 interventions should prioritise top-*K* cases scheduled within the critical 0–7 day lead-time bucket, where there is minimal time to reallocate theatre capacity. A minimum bundle should include rapid phone confirmation, identification of practical barriers (e.g., illness, transport, work/caring constraints), and an immediate rescheduling pathway when attendance is uncertain.

Tier 2 interventions prioritise top-*K* cases within 8–14 days and/or service lines with higher LCDNA burden, where teams have more time to act but early confirmation can still prevent late disruption. The workflow can be lighter-touch (e.g., earlier messaging/confirmation, completion of outstanding pre-operative steps, and contingency planning). Tier 3 uses the model as an audit tool: when a pathway or service line receives high model attribution but observed LCDNA rates are lower than expected, the mismatch should trigger pathway review—either existing processes are effectively mitigating cancellations or unmeasured operational factors are driving false positives. Using these discrepancies for audit supports continuous quality improvement rather than rigid rule-making.

These tiers are intended as practical implementation guidance. Trusts should adapt the exact intervention bundle and lead-time thresholds to local staffing and pathways, while retaining the core principle that near-term high-risk cases warrant the most consistent, capacity-protected response.

### 5.6. Interpretability and plausibility of learned signals

Explainability analyses using complementary global views (XGBoost gain-based importance, permutation importance under the operational objective, and SHAP) consistently identify lead-time, specialty indicators, patient history (e.g., cancellation rate), and calendar effects as dominant predictors. The convergence across these views increases confidence that model behaviour is not an artefact of a single importance metric. The SHAP patterns are also operationally plausible: shorter booking-to-procedure intervals are associated with higher predicted LCDNA risk, and patient-history features increase predicted risk for repeat cancellers. Day-of-week and holiday proximity effects indicate additional operational and behavioural structure that would be difficult to capture with manual heuristics.

These explanations should be read as associative rather than causal. Even so, they serve two practical purposes: (i) they help build stakeholder confidence by showing that the model relies on clinically and operationally interpretable signals, and (ii) they inform workflow design by indicating which types of cases are likely to dominate risk. The grouped importance view (Figure 9) is also easier to communicate to clinical stakeholders because it reduces fragmentation from one-hot encodings and summarises attribution at a category level.

### 5.7. Equity and fairness safeguards for deployment

Because the policy allocates limited human support, it should be deployed with explicit equity safeguards. While the primary objective here is operational value, the explanation analyses suggest that operational factors (lead time, service line, calendar effects, and prior attendance/cancellation history) dominate the learned signals, with demographic variables such as age and gender playing a secondary role. That is somewhat reassuring, but it does not remove the need for formal auditing.

We recommend embedding a routine fairness audit alongside performance monitoring: report Cost@K, recall@K, and missed-LCDNA rates by age band, gender, and deprivation/postcode-derived proxies where available; examine whether the top-*K* list disproportionately concentrates outreach on or away from any subgroup; and pre-define escalation thresholds (e.g., meaningful gaps in recall@K across groups) that trigger recalibration, workflow review, or subgroup-specific decision rules.

As an immediate priority, audits should cover the highest-volume patient cohorts in the dataset (as reflected in the descriptive distributions) to ensure that Trust-wide roll-out improves efficiency without introducing systematic disparities.

### 5.8. Positioning relative to prior work

Most prior cancellation-prediction studies focus on outpatient appointments or specific paediatric surgical contexts and typically report AUROC or accuracy as primary objectives Tuan et al. (2025); Liu et al. (2019); Luo et al. (2020); Zhang et al. (2021); Sardesai et al. (2024). Our study adds to that literature by focusing on NHS elective surgery LCDNA, explicitly optimising an operational cost objective, and evaluating under deployment-oriented regimes that include cross-hospital domain shift and time-forward testing. This helps bridge predictive modelling and theatre operations, where the real decision is how to allocate limited intervention resources to reduce avoidable disruption. Unlike prior studies that relied on internal record-level random splits within a single setting, we also enforce strict patient-level exclusivity so that no patient appears in both training and evaluation data. That reduces leakage through memorisation of patient-specific attendance traits and gives a more realistic estimate of performance and cost savings on unseen patients at deployment.

### 5.9. Limitations and future work

This study has several limitations. First, the dataset is drawn from a single Trust and a limited time period; although we explicitly test both temporal drift and cross-site operational shift, broader external validation across additional Trusts is still needed. Second, the analysis is observational: predicted savings depend on the assumed cost model and on the effectiveness of the chosen intervention bundle, which should ultimately be assessed prospectively. Third, local pathways and capacity constraints may change over time, requiring ongoing monitoring, recalibration, and governance.

Future work should include: (i) multi-Trust external validation and transfer learning strategies to support safe scaling; (ii) prospective or pragmatic implementation studies that measure realised cancellations avoided, staff time, and theatre utilisation; (iii) formal subgroup fairness evaluation integrated with the routine audits proposed above; and (iv) further refinement of specialty- and pathway-specific workflows informed by the tiered intervention framework and pathway-audit signals.

## 6. Conclusion

We present a decision-focused framework for reducing elective surgery LCDNA events by combining cost-sensitive risk ranking with a capacity-aware top-*K* intervention policy. Across two deployment-relevant validation regimes, the policy yields substantial expected-cost reductions and provides a practical basis for Trust-wide cancellation mitigation.

The dual-regime evaluation distinguishes temporal drift from cross-site operational shift and helps frame deployment expectations: cross-site performance offers a conservative baseline, while within-site time-forward performance provides an optimisation target. The proposed domain-shift risk budget and phased roll-out guidance translate these results into concrete implementation steps.

Explainability analyses highlight the importance of lead time and service-line pathways, and they help motivate tiered intervention workflows and pathway audits. Together with routine monitoring and embedded fairness checks, the approach is intended to deliver operational value while supporting safe, accountable deployment at scale.

## Data Availability

All data produced in the present study are available upon reasonable request to the authors and subject to NHS approval.

## 7. Key Findings for Policy Makers

This study shows that hospitals can reduce last-minute elective surgery cancellations by using data to identify patients who may need extra contact before surgery. The key findings are:

- **Targeted contact works better than contacting everyone or doing nothing.** Focusing on higher-risk patients reduced the expected cost per case from about £103 to £77 in the cross-site test.
- **Start cautiously and adapt locally.** The model may perform differently in different hospitals, so each site should start with a manageable approach and adjust it using local results.
- **The workload is practical.** In this study, the recommended approach would mean around 5 extra patient contacts per working day for a medium-sized hospital.
- **Focus first on patients close to their surgery date.** Patients due for surgery within the next 7 to 14 days should be prioritised, because there is less time to avoid wasted theatre slots.
- **The main risk signs are simple.** Higher risk is linked to short booking time, specialty or service line, previous cancellation history, and calendar effects such as day of week or holidays.
- **Use the model to improve services.** If some specialties or pathways often appear high-risk, managers should review whether booking, pre-assessment, or patient communication can be improved.
- **Monitor safety and fairness.** Hospitals should regularly check outcomes, staff workload, savings, and whether different patient groups are being treated fairly.

Overall, the model should be used as a practical support tool to help staff contact the right patients at the right time and reduce avoidable theatre disruption.

## Acknowledgments

The authors gratefully acknowledge Gloucestershire Hospitals NHS Foundation Trust for providing the dataset and operational context that made this research possible. We thank the University of Gloucestershire for supporting this collaborative research initiative. We also acknowledge the clinical and administrative staff at Gloucestershire Hospitals NHS Foundation Trust who provided valuable insights into the operational challenges of theatre utilisation and patient cancellations. This work was conducted as part of a collaborative research partnership between Gloucestershire Hospitals NHS Foundation Trust and the University of Gloucestershire, aimed at improving healthcare operational efficiency through data-driven approaches.

## Data Availability

The dataset used in this study contains sensitive information and operational data from Gloucestershire Hospitals NHS Foundation Trust. Due to privacy and confidentiality constraints, the raw dataset cannot be made publicly available. However, aggregated statistics and model performance metrics are reported in this paper. Researchers interested in accessing similar data should contact Gloucestershire Hospitals NHS Foundation Trust directly, subject to appropriate data governance and ethics approvals.

Source code and analysis scripts (including feature processing, split generation, model training, and evaluation under the cost–capacity policy) can be shared with reviewers and qualified researchers subject to NHS Trust data governance constraints and institutional approvals. A de-identified data dictionary, full feature list, and model specification (including hyperparameters) are available to support reproducibility, and the trained model artefacts can be disclosed under the same governance conditions where permitted.

## Ethics Approval and Consent to Participate

The study uses routinely collected healthcare data and was conducted under an approved analysis plan for a retrospective observational study; no separate prospective registration was undertaken. No formal patient and public involvement group was used in the design or conduct of the study. REDACTED has approved this research with the approval code: SREC.2025. 0217-1031. Individual patient consent was not sought, as the ethics approval and associated governance arrangements covered use of de-identified routine data and waived direct consent requirements. All eligible records in the study period were included, so sample size was determined by data availability rather than prospective power calculation; the resulting number of LCDNA events was sufficient for the planned model development and evaluation. Formal subgroup fairness testing was not the primary objective of this paper and is planned for prospective monitoring alongside routine audit. In a live deployment, missing or invalid predictor values would be handled by the preprocessing pipeline using leakage-safe feature gating and median imputation for numeric inputs, with structurally invalid records flagged for manual review rather than automated action.

## Appendix A. Supplementary Material

**Table A.13:** Supplementary comparison of TRAIN/CAL/VAL/TEST splits on key variables. Missingness is the share of rows with any missing value among age_at_admission, gender_original, specialty, or booking-to-procedure lead time.

| Split | N | Age (mean ± SD) | Female (%) | Top specialty (%) | Lead time, median [IQR] | GRH (%) | LCDNA (%) | Missingness (%) |
| --- | --- | --- | --- | --- | --- | --- | --- | --- |
| TRAIN | 13,040 | 62.0 ± 19.1 | 54.2 | Trauma and Orthopaedics (31.8) | 26 [16–36] | 0.0 | 10.85 | 0.0 |
| CAL | 2,348 | 46.6 ± 24.6 | 55.9 | ENT (33.8) | 22 [14–34] | 100.0 | 11.54 | 0.0 |
| VAL | 2,239 | 47.3 ± 24.8 | 56.0 | ENT (29.7) | 22 [13–34] | 100.0 | 10.99 | 0.0 |
| TEST | 2,142 | 46.0 ± 24.7 | 54.2 | ENT (33.1) | 24 [14–35] | 100.0 | 10.32 | 0.0 |

## Appendix B. Feature List

This appendix provides a complete list of features excluded for leakage prevention and features included in the final model.

### Appendix B.1. Features Excluded for Leakage Prevention

The following features were excluded because they contain information that would only be available after the procedure outcome is known, or they directly encode the target variable. These columns appear before the safe feature gate (procedure_year) in the dataset schema.

- `Actual_Procedure_Time_Min`
- `All_Procedures_Performed`
- `All_Procedures_Performed_OPCS_Codes`
- `Anaestetic_Performed`
- `Anaestetic_Performed_Code`
- `Anasethetic_Planned`
- `Anasethetic_Planned_Code`
- `Cancelled_Case`
- `IS_COPD`
- `IS_asthma`
- `IS_cancer`
- `IS_chronic_kidney_disease`
- `IS_coronary_heart_disease`
- `IS_dementia`
- `IS_diabetes`
- `IS_frailty_proxy`
- `IS_hypertension`
- `IS_mental_health`
- `OTD_Cancellation`
- `Planned_Theatre_Time`
- `Postcode`
- `Procedure Booked`
- `Procedure_Booked_Code`
- `Short_NOtice_Cancellation_<=3days`
- `Surgeon_Booked`
- `Surgeon_Booked_code`
- `Surgeon_Operated`
- `Surgeon_Operated_Code`
- `Theatre_Procedure_Performed`
- `Theatre_Procedure_Performed_Code`
- `Theatre_Suite_Booked`
- `Theatre_Suite_Booked_Code`
- `Theatre_Suite_Procedure_Performed`
- `Theatre_Suite_Procedure_Performed_Code`
- `age_at_admission`
- `anaesthestist`
- `anaesthestist_code`
- `booking_date`
- `cancellation_reason_freetext`
- `cancer_type`
- `derived_cancellation_date`
- `derived_cancellation_group1`
- `derived_cancellation_group2`
- `derived_cancellation_reason`
- `derived_cancellation_reason_code`
- `derived_cancellation_source`
- `diabetes_type`
- `ethnicity`
- `procedure_start_date`
- `religion`
- `specialty`
- `theatre_cancellation`
- `theatre_cancellation_code`
- `theatre_cancellation_date`

**Total excluded:** 54 features.

### Appendix B.2. Features Included in the Model

The following features were included in the final model. Features are listed in the order they appear in the dataset schema (starting from procedure_year onwards), excluding metadata columns (patient identifier, hospital, procedure date, target) and blacklisted features.

#### Appendix B.2.1. Base Features from Dataset

The following base features were extracted from the dataset schema (starting from `procedure_year onwards`), after excluding metadata columns and blacklisted features:

- `CGH`
- `CIR`
- `Days From Public Holiday`
- `Days From School Holiday`
- `Days To Public Holiday`
- `Days To School Holiday`
- `Days between booking and procedure`
- `Days_From_Public_Holiday_bucket`
- `Days_To_Public_Holiday_bucket`
- `Days_booking_procedure_bucket`
- `Friday`
- `IS_COPD_binary`
- `IS_asthma_binary`
- `IS_cancer_binary`
- `IS_chronic_kidney_disease_binary`
- `IS_coronary_heart_disease_binary`
- `IS_dementia_binary`
- `IS_diabetes_binary`
- `IS_frailty_proxy_binary`
- `IS_mental_health_binary`
- `Monday`
- `STG`
- `Saturday`
- `School Holiday`
- `Sunday`
- `TWC`
- `Thursday`
- `Tuesday`
- `Wednesday`
- `gender`
- `patient_appearances_before`
- `patient_cancellation_rate`
- `patient_cancellations_before`
- `procedure_day`
- `procedure_month`
- `procedure_month_bucket_v1`
- `procedure_month_bucket_v2`
- `procedure_year`
- `speciality_Breast_Surgery`
- `speciality_Colorectal_Surgery`
- `speciality_ENT`
- `speciality_Gynaecological_Oncology`
- `speciality_Gynaecology`
- `speciality_Ophthalmology`
- `speciality_Oral_Surgery`
- `speciality_Spinal_Surgery`
- `speciality_Trauma_and_Orthopaedics`
- `speciality_Upper_Gastrointestinal_Surgery`
- `speciality_Urology`
- `speciality_Vascular_Surgery`

**Total base features:** 50.

#### Appendix B.2.2. Blacklisted Features

The following features were excluded via the feature blacklist (`configured via feature_blacklist_substrings`) to prevent geographic information leakage or other operational concerns:

- `Distance to theater miles`
- `postcode_GL1`
- `postcode_GL2`
- `postcode_GL3`
- `postcode_GL4`
- `postcode_GL5`
- `postcode_GL6`
- `postcode_GL7`
- `postcode_GL8`
- `postcode_GL9`
- `postcode_others`

**Total blacklisted:** 11.

### Appendix B.3. Summary

- Leakage columns excluded: 54
- Base features included: 50
- Engineered features added: 0
- Blacklisted features excluded: 11
- Total final features: 50

## Footnotes

1 Explainability was computed on the GRH validation split only (N=2,239, prevalence ≈ 0.11) to preserve the sealed TEST set for unbiased reporting.

